# Brain Age Gap Reduction Following Physical Exercise Mirrors Negative Symptom Improvement in Schizophrenia Spectrum Disorders

**DOI:** 10.1101/2025.01.08.24319554

**Authors:** Deniz Yilmaz, Sergi Papiol, Daniel Keeser, James H Cole, Berend Malchow, Andrea Schmitt, Peter Falkai, Isabel Maurus, Lukas Roell

**Author notes:** Corresponding author: Deniz Yilmaz, M.Sc. Department of Psychiatry and Psychotherapy University Hospital, LMU Munich, Nussbaumstrasse 7, 80336 Munich, Germany. Equal Contribution (Isabel Maurus, Lukas Roell).

## Abstract

Schizophrenia spectrum disorders (SSD) are associated with accelerated brain aging, reflected in an increased brain age gap. This gap serves as a biomarker, indicating poorer brain health, cognitive deficits, and greater severity in specific symptom domains. Physical exercise holds promise as an adjunct therapy to mitigate these deficits by potentially promoting brain recovery. However, the extent of overall improvements in brain health following exercise, along with their predictors and relationships to symptom clusters, are yet to be determined. This study examined the brain age gap metric as a quantitative indicator of brain recovery in response to physical exercise. To achieve this, we aggregated data from two randomized controlled trials, analyzing baseline (*n* = 134) and 3- or 6-month post-exercise (*n* = 46) data from individuals with SSD. Our findings revealed that patients with a higher baseline BMI demonstrated greater brain recovery, as evidenced by a reduced brain age gap post-exercise. Furthermore, changes in the brain age gap were associated with improvements in negative symptoms and cognition, suggesting that reductions in brain-predicted age may reflect symptom relief, particularly in domains beyond positive symptoms. These results underscore the importance of BMI in brain health, support using the brain age gap as a surrogate marker for tracking clinically relevant brain recovery, and highlight the need for stratified interventions and combined lifestyle modifications to enhance outcomes in SSD.

**Glossary:** *Schizophrenia spectrum disorders (SSD):* Mental health conditions characterized by psychosis, an alteration of the perception of reality. Cardinal symptoms include hallucinations (sensory perception not mirroring reality) and delusions (persistent beliefs that are not rooted in reality).

*Positive symptoms:* A symptom cluster of SSD including complaints that are distinctively present in the patiens: hallucinations, delusions, and thought disorder (disorganized thinking and speech).

*Negative symptoms:* A symptom cluster of SSD including complaints that are distinctively absent in the patiens: loss of interest, motivation, enjoyment, and social interactions, flattened affect.

*Cognitive symptoms:* Another cluster of symptoms in SSD including deficits in attention, executive function, and memory.

*Biomarker:* Objective, quantifiable indicators of biological states or processes used to predict, diagnose, and treat illnesses.

*Brain age gap:* A biomarker of brain health and aging. Brain-predicted age is the age predicted by a machine learning algorithm based on brain imaging data. Subtracting chronological age from the brain-predicted age results in the brain age gap, where positive values indicate an accelerated aging of the brain.

*Neuroplasticity:* The brain’s ability reorganize itself through new synaptic connections following learning, treatment, or injury.

*Randomized Controlled Trials (RCTs):* A study design that randomly assigns participants to an experimental group or a control group to test the efficacy of an intervention.

## 1. Introduction

Ample evidence suggests that schizophrenia spectrum disorders (SSD) are associated with both brain volume loss (Adriano et al., 2012; Hulshoff Pol et al., 2002; van Erp et al., 2016; van Haren et al., 2008) and faster brain aging compared to healthy controls (Kaufmann et al., 2019). Consistent with this, machine learning algorithms based on structural brain imaging predict that SSD patients have a higher brain-predicted age than their chronological age (Ballester et al., 2022; Constantinides et al., 2023; Hajek et al., 2019; McWhinney et al., 2021, 2022; Nenadić et al., 2017; Shahab et al., 2019; Wrigglesworth et al., 2021). This difference, or *brain age gap*, constitutes a data-driven biomarker of brain health, with larger gaps indicating accelerated brain aging. Such biomarkers are promising as they can serve as a singular but global neurobiological outcome metric in targeted clinical interventions and aid in medical monitoring. Additionally, they can function as predictive identifiers of disease progression and treatment response (Biondo et al., 2022a; Cole et al., 2018; Falkai et al., 2018; Fan et al., 2024; Liew et al., 2023; Lu et al., 2024; Tseng et al., 2024; Van Gestel et al., 2019)

The brain age gap correlates negatively with mindfulness and positively with rumination in schizophrenia but not in controls (Abram et al., 2023) and is associated with reduced working memory and processing speed (Wang et al., 2021). Moreover, faster brain aging in first episode psychosis (FEP) positively correlates with worsening negative symptoms and functioning (McWhinney et al., 2021), highlighting its clinical relevance. The neurodevelopmental hypothesis of schizophrenia provides a theoretical framework for these empirical findings (Fatemi and Folsom, 2009; Marenco and Weinberger, 2000; Murray and Lewis, 1987). It posits that excessive synaptic pruning during development leads to accelerated synapse loss, contributing to the brain age gap (Demro et al., 2022; Hua et al., 2024; Ling et al., 2024). This overpruning disrupts the excitation/inhibition balance, driving the negative and cognitive symptoms, while mesolimbic projections downstream from this imbalance are hypothesized to underlie dopaminergic alterations and positive symptoms (Howes and Shatalina, 2022). This hypothesis aligns with the strong genetic basis of SSD pathogenesis (Ling et al., 2024; Schmitt et al., 2023; Trubetskoy et al., 2022) and genetic modulation of treatment response (Papiol et al., 2024, 2019, 2017).

Along with the positive symptoms, which are often alleviated by antipsychotic medication, cognitive deficits, and negative symptoms are the cardinal features of SSD (Thiebes et al., 2017a). Despite the prevalence of the latter two, they remain difficult to treat, thereby encumbering the patients and contributing significantly to the global disease burden (Mulert, 2012; Thiebes et al., 2017b; Vos et al., 2020). As a promising add-on treatment, physical exercise has consistently shown beneficial effects in alleviating these cognitive and negative symptoms in addition to improving overall functioning (Dauwan et al., 2021, 2016, 2016; Falkai et al., 2017; Fernández-Abascal et al., 2021; Firth et al., 2016, 2015, 2015; Kim et al., 2023; Maurus et al., 2021, 2019a, 2019b; Sabe et al., 2020; Schmitt et al., 2018; Shimada et al., 2022; Vancampfort et al., 2015; Vogel et al., 2019; Xu et al., 2022).

However, the neural mechanisms driving improvements in cognitive impairment, negative symptoms, and overall functioning after physical exercise remain to be clarified. Current preliminary evidence in this field suggests that physical exercise as an add-on treatment for SSD may increase hippocampal volume (Girdler et al., 2019; Pajonk et al., 2010; but see Malchow et al., 2016), anterior left temporal gyrus volume (Malchow et al., 2016), white matter integrity in tracts related to motor functioning (Svatkova et al., 2015), and serum BDNF levels —a mediator of structural and functional neuroplasticity (Colucci-D’Amato et al., 2020; Knaepen et al., 2010). The treatment also counteracted the typically observed disturbances in the default-mode network, the cortico-striato-pallido-thalamo-cortical, and the cerebello-thalamo-cortical circuits in a group of SSD patients, who underwent a physical exercise intervention for 6 months (Roell et al., 2022). Furthermore, the right posterior cingulate gyrus volume increase was related to clinical improvements. These findings show that physical exercise has the potential to affect multiple clinically relevant brain regions and networks in SSD. Nonetheless, possible global brain health improvements after physical exercise and their link to the precise symptomatic changes over time are yet to be determined. Addressing this question is particularly relevant, as in healthy older adults, a reduced brain age gap relates strongly to physical activity, including daily habits like stair climbing (Bittner et al., 2021; Steffener et al., 2016), and correlates with current and retrospective physical fitness (Ouyang et al., 2022; Wing et al., 2024).

Here, we investigated the role of brain age as a singular measure of global brain health in exercise interventions for SSD, focusing on its sensitivity to capture clinical outcomes. First, we hypothesize that at baseline a larger positive brain age gap is associated with lower aerobic fitness, more severe clinical symptoms, and a higher genetic burden for SSD. Second, we assume that the brain age gap decreases following physical exercise treatment in SSD. Third, we expect that longitudinal decreases in the brain age gap are linked to improvements in clinical and cognitive symptoms. Fourth, we presume that the brain age gap at baseline predicts the amount of change in clinical and cognitive symptoms over time and vice versa. Lastly, we propose that a lower genetic burden for SSD predicts greater improvements in the brain age gap following the intervention. If our hypotheses are supported, the brain age gap variable could serve as a proxy for neurobiological improvements post-treatment and potentially aid in predicting treatment response, ultimately enhancing the prediction, monitoring, and quantification of outcomes.

## 2. Methods

### 2.1. Participants

#### 2.1.1. Test Set—Patient Cohort

We pooled the data of individuals with SSD (F20* diagnosis according to ICD-10, World Health Organization, 2004; confirmed by the Mini-International Neuropsychiatric Interview, Version 6.0.0, Sheehan et al., 1998) from two randomized controlled clinical trials with a similar design (Exercise2 and ESPRIT studies; for details see Malchow et al., 2016 and Maurus et al., 2021, registered at www.clinicaltrials.gov under NCT01776112 and NCT03466112, respectively). Inclusion criteria required participants to be on a stable regimen of antipsychotic medication. Six subjects from the ESPRIT study were excluded due to the low quality of their T1-weighted scans. At baseline, T1-weighted MRI scans of *n* = 134 (47 females, *M*_age_ = 36.44, *SD*_age_ = 12.49, ranged from 18-61, *n* = 40 from Exercise2 and *n* = 94 from ESPRIT) participants were available. Post-exercise data obtained from the Exercise2 study were collected following 3 months of regular exercise, whereas the ESPRIT sample undertook 6 months of regular exercise. Both studies were in accord with the Declaration of Helsinki and were approved by the local ethics committees. All participants gave written consent. See Tables 1, 2, and 3 for sample characteristics of the pooled data, supplementary materials for the exclusion criteria, and https://osf.io/tr3nx/ for further materials of ESPRIT and the current paper.

**Table 1.** Descriptive Statistics for the Data Used in Baseline Analysis.

| Characteristic | Female ($n = 47$)<sup>1</sup> | Male ($n = 87$)<sup>1</sup> | Overall ($n = 134$)<sup>1</sup> |
| --- | --- | --- | --- |
| Age | 38.77 (13.03) | 35.18 (12.08) | 36.44 (12.49) |
| Brain-Predicted Age | 34.20 (8.23) | 37.11 (11.72) | 36.09 (10.69) |
| Brain Age Gap | -4.57 (9.14) | 1.92 (7.73) | -0.35 (8.79) |
| Aerobic Fitness | 0.75 (0.28) | 0.94 (0.43) | 0.87 (0.39) |
| (Missing) | 13 | 24 | 37 |
| Years of Education | 14.97 (4.41) | 14.30 (3.75) | 14.54 (3.99) |
| CPZ | 496.38 (512.40) | 506.65 (363.50) | 503.05 (419.85) |
| Number of Trainings | 24.17 (19.15) | 28.47 (16.13) | 26.96 (17.30) |
| Body Mass Index | 27.94 (5.27) | 28.03 (4.76) | 28.00 (4.93) |
| PANSS Positive | 11.98 (5.21) | 12.02 (4.57) | 12.01 (4.79) |
| PANSS Negative | 14.09 (6.77) | 14.91 (7.14) | 14.62 (7.00) |
| PANSS General | 28.64 (11.03) | 28.83 (12.01) | 28.76 (11.63) |
| PANSS Total | 54.70 (20.98) | 55.76 (21.18) | 55.39 (21.04) |

<sup>1</sup>Mean (Standard Deviation)
*Note.* CPZ: Chlorpromazine-Equivalent Dose; PANSS: Positive and Negative Syndrome Scale; DST WM: Digit Span Test Working Memory; TMT: Trail Making Task; VLMT: Verbal Learning and Memory Test; STM: Short Term Memory; LMT: Long Term Memory.

**Table 2.** Descriptive Statistics and Bayes Factors for the Data Used in the Longitudinal Analysis.

| Characteristic | Baseline ($n = 46$)<sup>1</sup> | Post-Exercise ($n = 46$)<sup>1</sup> | BF<sub>10</sub> |
| --- | --- | --- | --- |
| Age | 36.15 (11.41) | 36.15 (11.41) | - |
| Brain-Predicted Age | 36.43 (9.11) | 36.38 (9.37) | 0.22 |
| Brain Age Gap | 0.27 (9.46) | 0.22 (9.52) | 0.22 |
| Aerobic Fitness | 0.90 (0.42) | 1.09 (0.46) | 0.30 |
| (Missing) | 8 | 20 |  |
| Years of Education | 15.11 (4.39) | 15.11 (4.39) | - |
| CPZ | 591.03 (564.21) | 591.03 (564.21) | - |
| Number of Trainings | - | 36.54 (13.35) | - |
| Body Mass Index | 29.04 (4.79) | 28.28 (3.18) | 0.42 |
| PANSS Positive | 11.59 (5.78) | 10.85 (4.69) | 0.50 |
| PANSS Negative | 14.59 (7.33) | 13.24 (6.22) | 0.86 |
| PANSS General | 27.02 (13.22) | 25.50 (10.97) | 0.52 |
| PANSS Total | 53.20 (24.32) | 49.61 (20.10) | 0.85 |

<sup>1</sup>Mean (Standard Deviation)
*Note.* CPZ: Chlorpromazine-Equivalent Dose; PANSS: Positive and Negative Syndrome Scale; DST WM: Digit Span Test Working Memory; TMT: Trail Making Task; VLMT: Verbal Learning and Memory Test; STM: Short Term Memory; LTM: Long Term Memory. Jeffreys' Bayes factor, BF<sub>10</sub>, indicates the data-supported evidence for the alternative hypothesis that there is a difference between pre and post sessions. There was no evidence for such a difference for any of the variables. Wherever BF<sub>10</sub> is marked with “-”, the pre and post values were identical, except for Number of Trainings, which could be extracted only post-exercise.

**Table 3.**
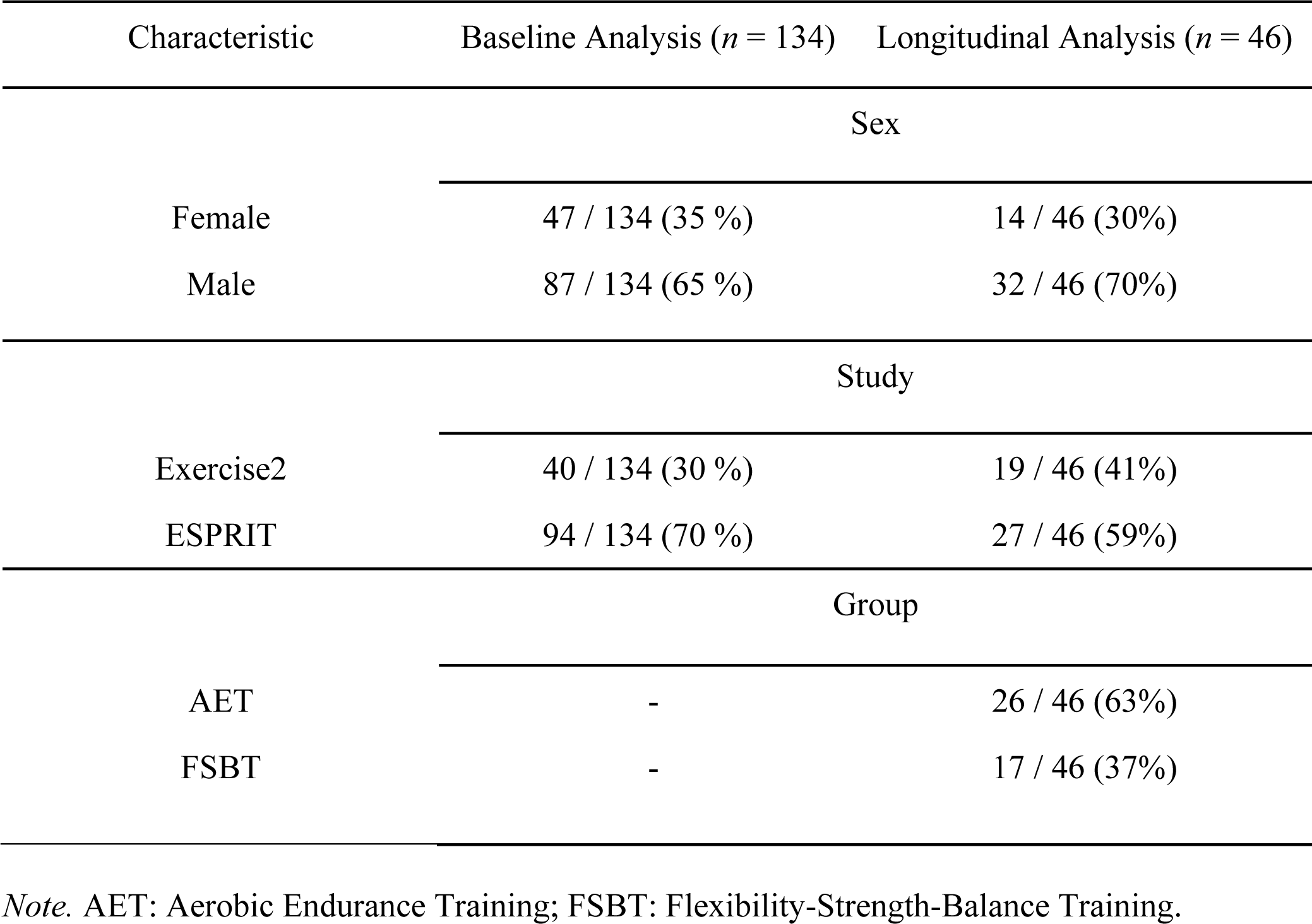
Frequencies of the Categorical Data.

| Characteristic | Baseline Analysis ($n = 134$) | Longitudinal Analysis ($n = 46$) |
| --- | --- | --- |
| Sex |  |  |
| Female | 47 / 134 (35 %) | 14 / 46 (30%) |
| Male | 87 / 134 (65 %) | 32 / 46 (70%) |
| Study |  |  |
| Exercise2 | 40 / 134 (30 %) | 19 / 46 (41%) |
| ESPRIT | 94 / 134 (70 %) | 27 / 46 (59%) |
| Group |  |  |
| AET | - | 26 / 46 (63%) |
| FSBT | - | 17 / 46 (37%) |

*Note.* AET: Aerobic Endurance Training; FSBT: Flexibility-Strength-Balance Training.

#### 2.1.2. Training Set—Healthy Cohort

To calculate the brain-predicted ages of the patient cohort, we used the open-access validated brainageR model (Biondo et al., 2022b; Clausen et al., 2022; Dörfel et al., 2023; Hobday et al., 2022), trained on T1-weighted MRI scans of *n* = 3377 healthy individuals from seven publicly available datasets (*M*_age_ = 40.6 years, *SD*_age_ = 21.4, ranged 18-92) and tested on a held-out set of *n* = 857 (*M*_age_ = 40.1 years, *SD*_age_ = 21.8, ranged 18-90). See https://github.com/james-cole/brainageR for further information about the healthy cohort characteristics.

### 2.2. Materials & Measures

#### 2.2.1. BrainAGE Prediction Methods

The brainageR software (v2.1) segments and normalizes the raw T1-weighted scans with SPM12 (Penny et al., 2011) and quality controls using the FSL *slicesdir* function. This ensures identical pre-processing of the healthy controls’ training data and the patients’ test dataset. It uses a Gaussian Processes Regression implemented in R to predict the brain age of individuals based on their combined vectorized and masked (0.3 probability threshold from the average image template derived from the brainageR model training set) grey matter, white matter, and cerebrospinal fluid images. We converted the dockerized version of brainageR (Prados, 2024) into a singularity container (available at https://osf.io/9jmrt/) and executed it with the data collected from the patients, yielding the brain-predicted ages of each patient. The brain age gap was then calculated by subtracting chronological age from the brain-predicted age per participant (see Figure 1). A greater brain age gap, therefore, indicates that the algorithm predicted the person to be older than their chronological age based on their neuroimaging data, suggesting that they have previously experienced accelerated brain aging. BrainageR demonstrated very high test-retest reliability, with an intraclass correlation coefficient (ICC) of 0.98 (95% CI [0.98–0.99]) and an adjusted mean absolute difference (Adj MAD) of 1.27 years within a year (Dörfel et al., 2023), making it an optimal choice for our longitudinal design. In the held-out test sample, Pearson’s correlation coefficient of *r* = 0.973 was found between chronological age and brain-predicted age (mean absolute error [MAE] = 3.933 years, range = 18-90 years, *n* = 857). In external validation, the model maintained high performance (*r* = 0.947, MAE = 4.90 years, 18-90 years, *n* = 611). Further details of this sample and model have been explained elsewhere (https://github.com/james-cole/brainageR) (Cole, 2019; Biondo et al., 2022; Clausen et al., 2022; Hobday et al., 2022).

**Figure 1.**
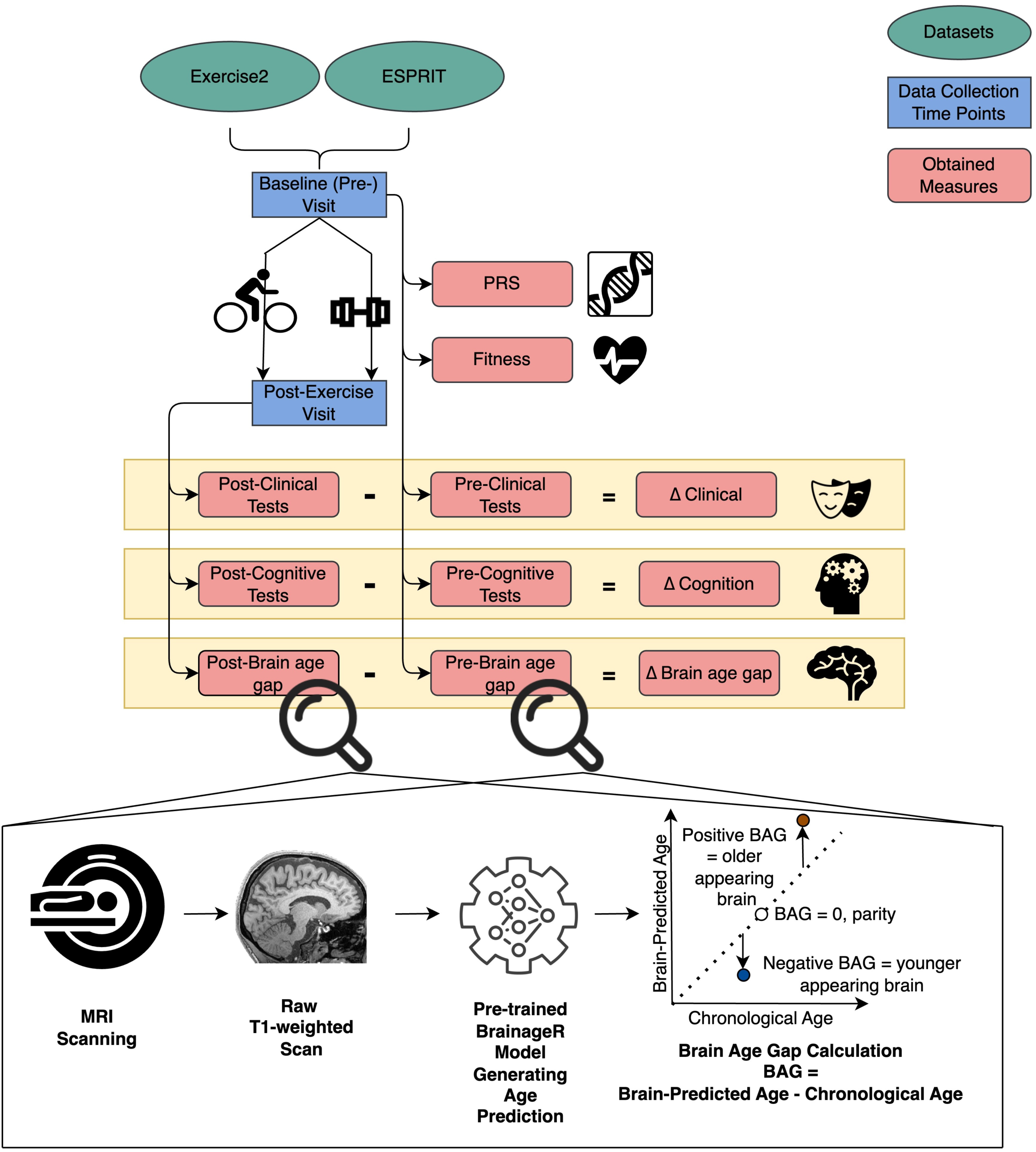
Study Protocols and Data Processing Workflow. *Note.* The workflow schematic includes retrieving the Exercise2 and ESPRIT datasets, comprising baseline (pre-exercise) and 3- or 6-month post-exercise data collection time points. The participants were either in the Aerobic Exercise Training group or Flexibility Strength and Balance group, depicted with a bicycle and dumbbell, respectively. Other than PRS, which was calculated only from the baseline blood samples, and fitness, for which only baseline scores were utilized in this paper, all measures were assessed both pre- and post-exercise. Change values for these variables were calculated by subtracting the post- from the pre-exercise value. The brain age gap was calculated with the identical procedure using pre- and post-exercise T1-weighted MRI scans. This procedure involves processing raw MRI data using the brainageR software, which generates brain-predicted ages based solely on the MRI data. The brain age gap is then calculated by subtracting each participant’s chronological age from their brain-predicted age. BAG: brain age gap; Δ: change; PRS: schizophrenia polygenic risk score.

#### 2.2.2. Fitness Measures

The exercise intensity at which blood lactate concentration reaches 2 mmol/l denotes the aerobic threshold (Faude et al., 2009). Based on this, both studies assessed aerobic fitness using the ergometer-based lactate threshold test, identifying the individual aerobic threshold at a lactate concentration of 2 mmol/l (as elaborated by Maurus et al., 2021 in their exercise protocol). The wattage reached at this threshold was standardized by dividing it by body weight, yielding the aerobic fitness score per participant, representing the capacity to perform at an aerobic exercise intensity. Increasing scores indicate better fitness.

#### 2.2.3. Clinical Measures

The positive, negative, and total symptom severity was assessed with the Positive and Negative Syndrome Scale (PANSS; Kay et al., 1987), yielding the variables PANSS-positive, PANSS-negative, PANSS-general, and PANSS-total, the latter two indicating general non-SSD-specific psychopathology and the summarized symptom severity from the three subscales, respectively. Increasing PANSS scores denote more severe symptoms. The general functioning level was assessed using the Global Assessment of Functioning scale (GAF; Endicott et al., 1976). We calculated the chlorpromazine-equivalent medication dose (CPZ, for short) for each participant from their current antipsychotic medication (Leucht et al., 2016).

#### 2.2.4. Cognitive Measures

Trail Making Tests A and B (TMT mean denoting their standardized average; Reitan, 1985) assessed *global cognition* encompassing visual search, cognitive flexibility, processing speed, scanning, and executive functions; the backward Digit Span Test (DST; Tewes, 1994) measured *verbal working memory* performance; and the short-term and long-term components of the Verbal Learning and Memory Test (VLMT; Helmstaedter et al., 2009) evaluated *verbal declarative memory*. The *composite cognitive score* denotes the normalized average of all the aforementioned constructs. Higher scores indicate greater performance for all variables (see supplementary materials for the score calculations).

#### 2.2.5. Polygenic Risk Scores

Genotyping, quality control, and imputation were conducted as described elsewhere (Boudriot et al., 2024). Polygenic risk scores (PRS) were calculated using the PRS-CS tool, based on the most recent schizophrenia genome-wide association study of the Psychiatric Genomics Consortium (Trubetskoy et al., 2022). A higher PRS indicates an increased genetic burden for schizophrenia.

### 2.3. Procedure

Both studies had the following procedures: screening, baseline data collection, randomization into a treatment arm, three (Exercise2) or six (ESPRIT) months of physical exercise intervention, and post-exercise treatment data collection. Exercise2 included one exercise intervention arm of Aerobic Endurance Training with add-on computer-assisted cognitive remediation (AET and CACR; AET on bicycle ergometers, the full procedure explained by Malchow et al., 2016). Whereas, ESPRIT had two exercise arms: AET and Flexibility-Strength- Balance Training (FSBT; for further details see Maurus et al., 2021). Exercise2 and ESPRIT studies were overall very similar except for the double duration and the additional active arm of the ESPRIT protocol, and the addition of a 6-week CACR in Exercise2. We used all the acquired baseline data from SSD patients for the baseline analysis. For the longitudinal analysis, we only used the data from patients, who completed an exercise intervention including the pre- and post-exercise MRI measurements. Figure 1 depicts the study protocols and data processing workflow.

### 2.4. Statistical Analysis

We based our inferences on Jeffreys’ Bayes factor (BF_10_), a continuous quantification of the data-supported evidence for the alternative hypothesis (H_1_: *r* ≠ 0) relative to the null hypothesis (H_0_: *r* = 0). A BF_10_ of 1.5, for instance, would indicate that the given data is 1.5 times more likely to be observed under the alternative hypothesis. We employed the conventional framework to infer different strengths of evidence (1<BF_10_<3: anecdotal evidence; 3<BF_10_<10: moderate evidence; 10<BF_10_<30: strong evidence; BF_10_>30: very strong evidence for the alternative hypothesis) as well as examining the probability of direction (PD) and the region of practical equivalence (ROPE; for a more detailed explanation, see Makowski et al., 2019). For all coefficients in the regression models, we applied a conservative Gaussian prior (*M* = 0, *SD* = 1), reducing the likelihood of extreme values. A medium-narrow prior was used for the correlation analysis, prioritizing smaller effects and rendering large effects less probable. Thereby, we aimed to allow for modest effects and avoid overfitting.

We computed the brain age gap scores from brainageR as outlined above. Since the model does not automatically correct the predictions for the statistical dependency on chronological age, we included age as a covariate in all analyses. After calculating the brain age gaps, all analyses were conducted on R v4.4.1(2022). We opted for Pearson correlation for all correlation analyses except when assumptions were violated, in which case we ran Spearman correlations. For the categorical variables of sex (female vs. male), study (Exercise2 vs. ESPRIT), session (baseline vs. post-exercise), and group (AET vs. FSBT), we created dummy variables, with the first (leftmost) level in each comparison designated as the reference. We imputed missing values using k-nearest neighbors (kNN, *k* = 5) imputation if at least 80% of a given variable was available (see supplementary materials). Our analyses included outliers to account for the full spectrum of natural variability in behavior and physiology.

To test our first hypothesis regarding the association between baseline brain age gap on the one hand and aerobic fitness levels and clinical parameters on the other hand, we ran Bayesian partial correlations adjusted for the baseline variables of age, sex, CPZ, education years, BMI, and study. To examine our second hypothesis that session impacts the brain age gap longitudinally in the context of physical exercise, we used a Bayesian linear mixed-effects model with the session as a within-subjects factor, covariables as fixed effects, and random intercepts and slopes for subjects:

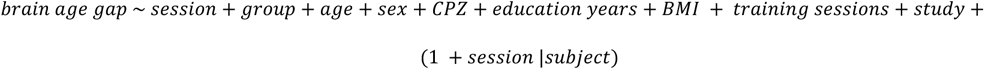

We calculated the change scores for PANSS, cognition, and brain age gap by subtracting baseline scores from those obtained after the exercise intervention (see Figure 1). Thus, a negative change score in PANSS or positive change score in cognition indicates symptom improvement, while a negative change in the brain age gap signifies a younger predicted brain age post-exercise interpreted as brain recovery. To test our third hypothesis regarding the relationship between changes in the brain age gap and improvements in clinical and cognitive parameters, we used Bayesian partial correlations, adjusting for age, sex, CPZ, education years, baseline BMI, number of training sessions, exercise group, and study. Additionally, our fourth hypothesis examined the potential predictive identifiers of treatment response. To this end, we ran Bayesian partial correlations between the baseline brain age gap and subsequent clinical changes, and vice versa— testing whether baseline clinical scores predict changes in the brain age gap. Similarly, we explored whether demographic variables and covariates (age, education years, baseline BMI, CPZ, and number of training sessions) were related to brain age gap change with Bayesian partial correlations. We performed an analogous analysis to evaluate our last hypothesis on the association between schizophrenia polygenic risk scores and changes in the brain age gap.

## 3. Results

Descriptive statistics for the data used in the baseline and longitudinal analyses are reported in Tables 1, 2, and 3.

### 3.1. Association between Baseline Brain Age Gap, Aerobic Fitness, Clinical Measures, and PRS

Baseline brain age gap was not associated with fitness, symptom severity, cognitive impairment, or PRS scores.

### 3.2. Longitudinal Changes Following Physical Exercise Intervention

To examine the longitudinal changes in the brain age gap in the context of physical exercise, we ran a Bayesian linear mixed-effects model, which showed no evidence of an effect of the session, indicating no overall change in the brain age gap over time post-exercise (β = 0.13, 95% CI [-0.44, 0.71], BF_10_ = 0.31; see Figure 2). However, a secondary finding from this model provided very strong evidence of the effect of BMI on the brain age gap (β = 0.26, 95% CI [0.12, 0.39], BF_10_ = 134.81), demonstrating that a lower BMI was associated with a smaller brain age gap.

**Figure 2.**
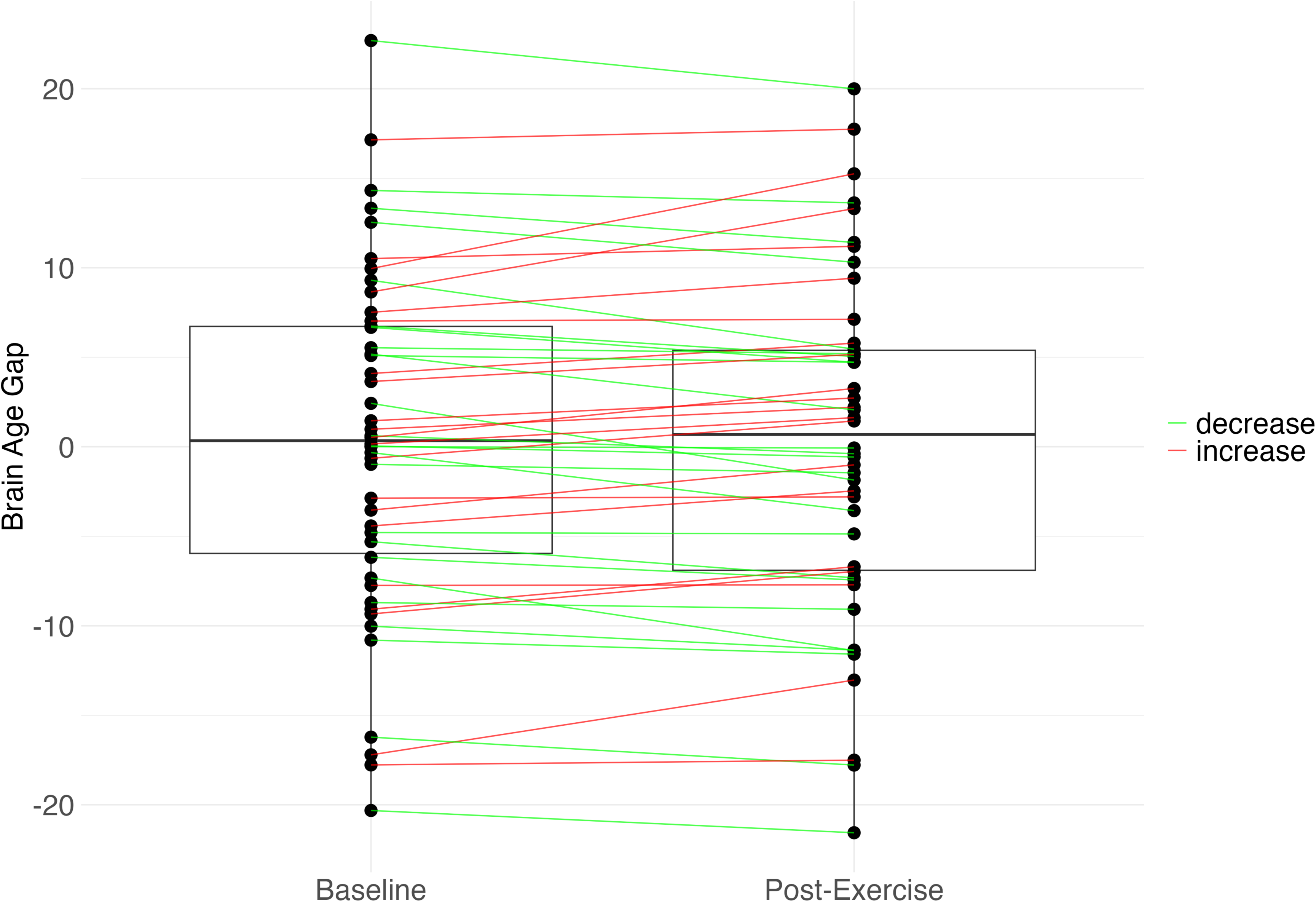
Session Effects on Brain Age Gap. *Note*. Data aggregated from the Exercise2 and ESPRIT datasets. Boxplots represent the brain age gap at baseline and post-exercise sessions. Individual participants are depicted as dots, with lines connecting them to illustrate change. Participants showing brain recovery (a decreased brain age gap) are connected to their post-exercise session in green, while those with an increased brain age gap are shown in red.

A partial Spearman rank correlation revealed moderate evidence of a positive relationship between brain age gap change and PANSS negative change (*r* = 0.27, 95% CI [0.02, 0.50], BF_10_ = 3.22, PD = 0.98, ROPE = 0.10), indicating that post-exercise brain recovery is linked to negative symptom improvement. PANSS positive, general, and total change were not related to brain age gap change (see Figure 3).

**Figure 3.**
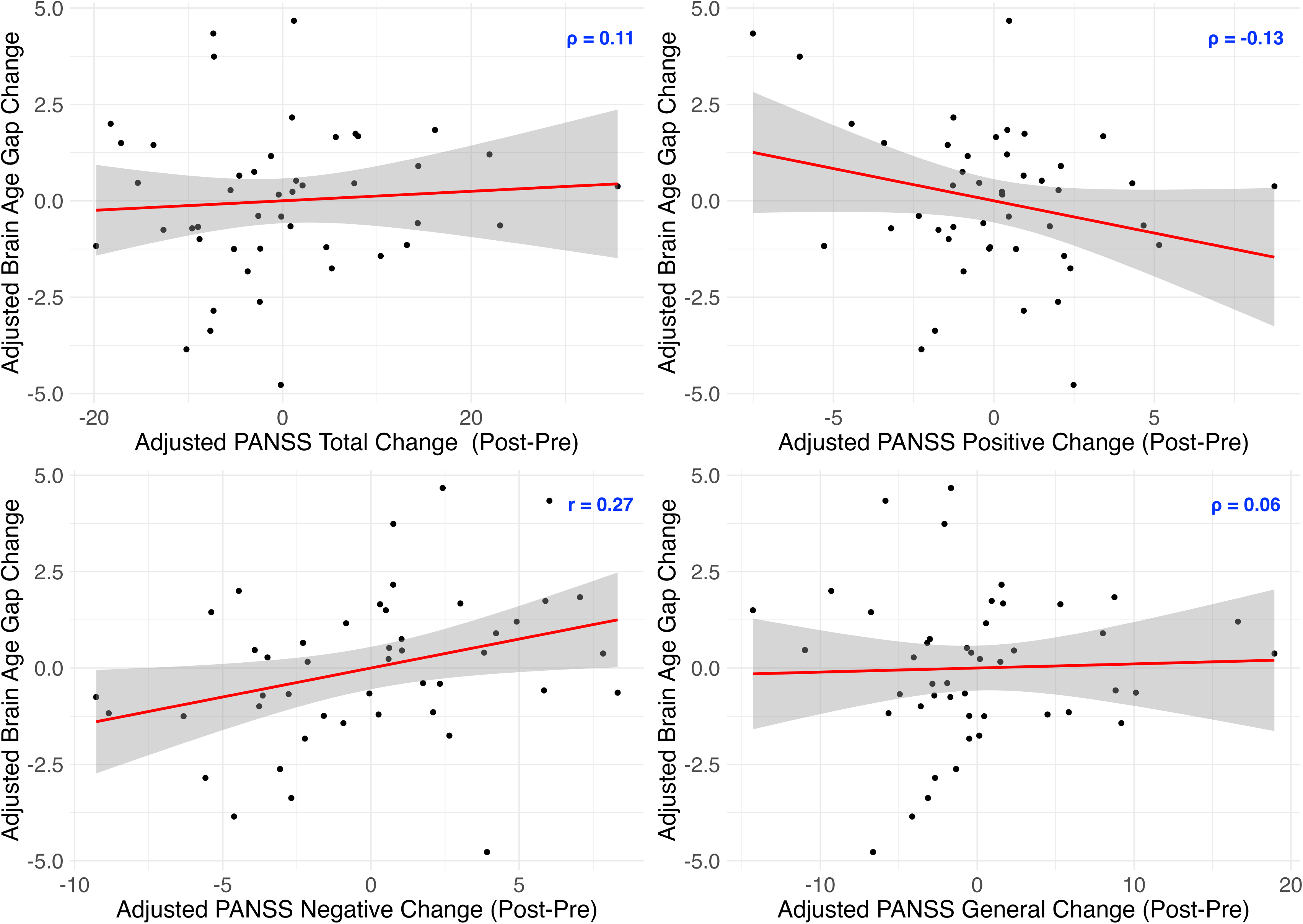
Partial Correlation Plots: Post-Exercise Brain Age Gap Change vs. PANSS Change. *Note.* The graphs display the partial correlation between brain age gap change (post-pre) and PANSS total change, as well as its subscales: positive, negative, and general change. Change is operationalized as Post-Pre for all variables. Therefore, a higher positive brain age gap change indicates greater brain aging and a higher positive PANSS change reflects increased symptom severity. Conversely, negative changes indicate a reduction in both brain aging and symptom severity. All variables are adjusted for covariates, and thus, the values shown represent residuals.

We found anecdotal evidence of a negative relationship between brain age gap change and a change in composite cognitive score (ρ = -0.26, 95% CI [-0.50, -0.01], BF_10_ = 2.60, PD = 0.97, ROPE = 0.11) and VLMT long-term memory (*r* = -0.25, 95% CI [-0.49, -0.01], BF_10_ = 2.36, PD = 0.97, ROPE = 0.12). Hence, brain recovery post-exercise was related to improvements in general cognition and long-term memory. There was no evidence that brain age gap change is associated with a change in TMT mean, DST working memory, or VLMT short-term memory (see Figure 4).

**Figure 4.**
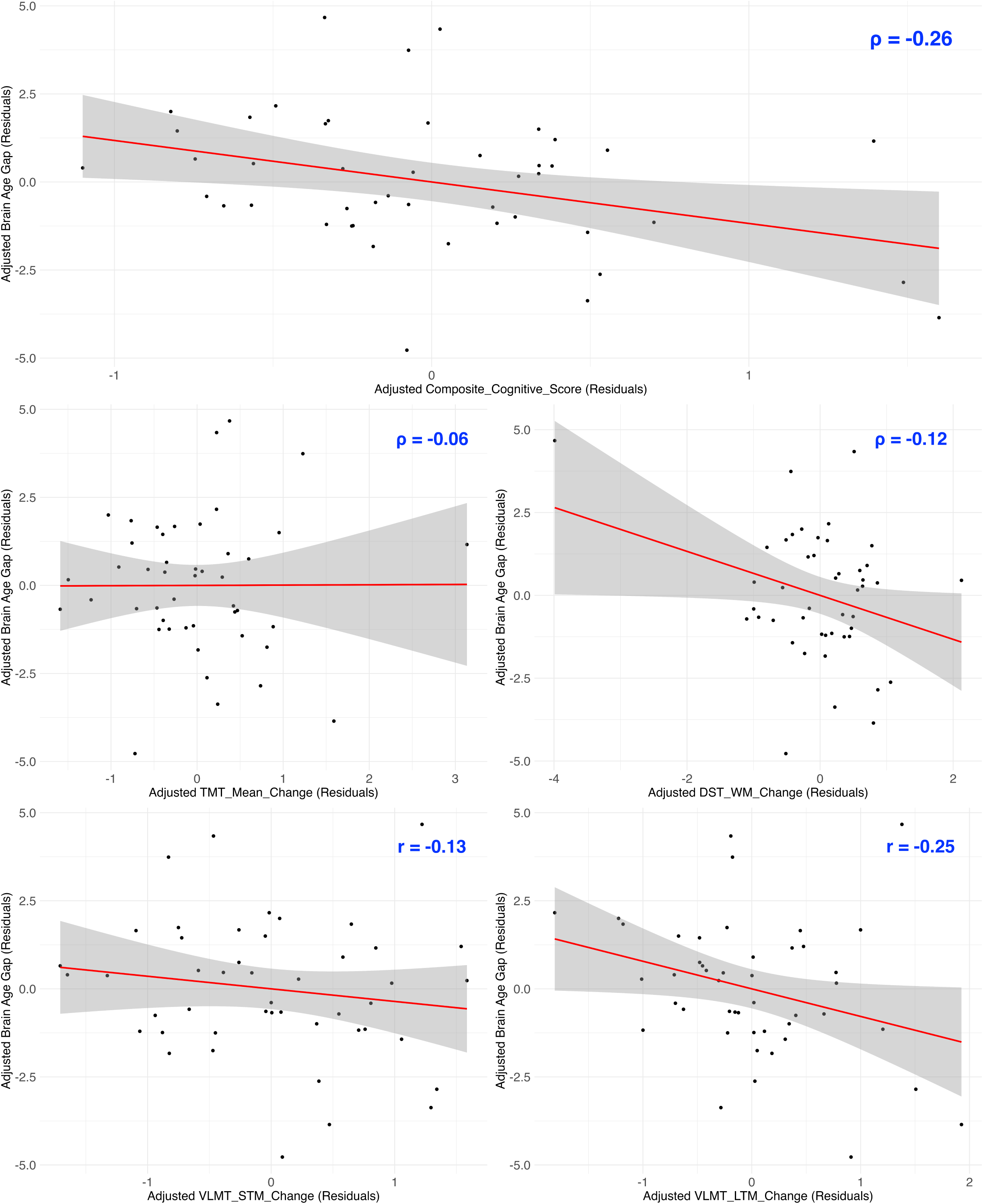
Partial Correlation Plots: Post-Exercise Brain Age Gap Change vs. Cognitive Change. *Note.* The graphs display the partial correlation between brain age gap change (post-pre) and composite cognition change, as well as change in its subscales: TMT mean, DST working memory (DST WM), VLMT short-term memory (VLMT STM), and VLMT long-term memory (VLMT LTM). Change is operationalized as Post-Pre for all variables. Therefore, a higher positive brain age gap change indicates greater brain aging but a higher positive cognitive change reflects improved cognition. Conversely, negative changes indicate a reduction in brain aging but a worsening of cognition. All variables are adjusted for covariates, and thus, the values shown represent residuals.

We inspected the predictors of treatment response and found that the baseline brain age gap was not associated with subsequent changes in the PANSS total, PANSS positive, PANSS negative, or PANSS general. However, examination of cognitive domain revealed anecdotal evidence suggesting that a lower brain age gap at baseline predicts greater subsequent improvement in composite cognitive score (ρ = -0.22, 95% CI [-0.47, 0.02], BF_10_ = 1.71, PD = 0.95, ROPE = 0.17), TMT mean (ρ = -0.23, 95% CI [-0.47, 0.02], BF_10_ = 1.80, PD = 0.96, ROPE = 0.15), and VLMT long-term memory (*r* = -0.21, 95% CI [-0.44, 0.06], BF_10_ = 1.39, PD = 0.94, ROPE = 0.20). No evidence was found for DST working memory change or VLMT short-term memory change (see Figure 5).

**Figure 5.**
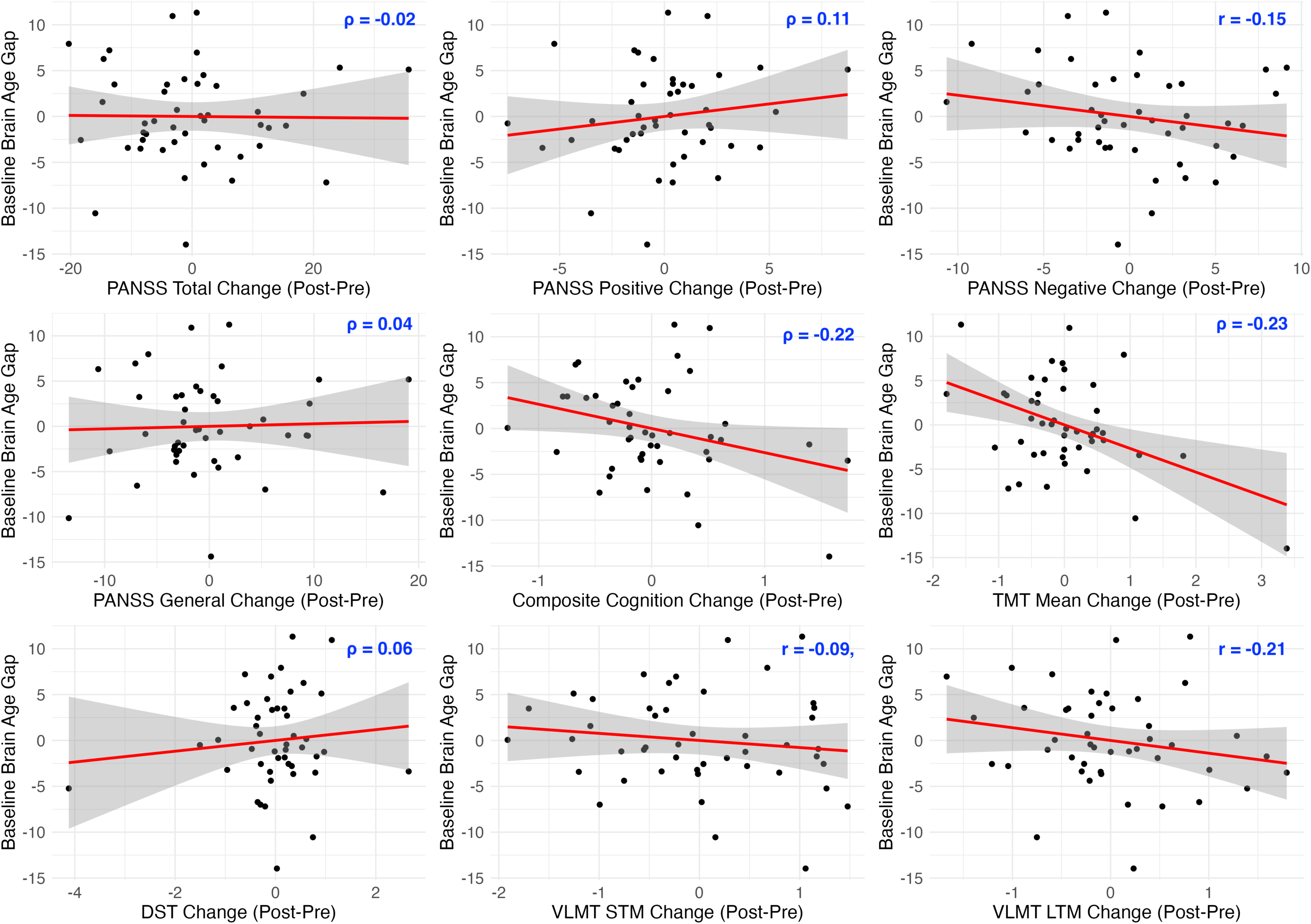
Partial Correlations: Brain Age Gap at Baseline vs. Clinical Change (Adjusted Values) *Note.* The graphs display the partial correlations between the baseline brain age gap and change in all clinical and cognitive variables. Change is operationalized as post-pre for all variables. Therefore, higher PANSS change values reflect increased symptom severity after the intervention, whereas, for cognitive variables, higher change values indicate improved cognition. All variables are adjusted for covariates, and thus, the values shown represent residuals. STM: short-term memory; LTM: long-term memory.

The relationship in the opposite direction, concerning the predictive value of baseline clinical characteristics on subsequent brain age change was observed anecdotally for baseline PANSS total (ρ = 0.17, 95% CI [-0.08, 0.42], BF_10_ = 1.01, PD = 0.91, ROPE = 0.26), PANSS positive (ρ = 0.20, 95% CI [-0.07, 0.43], BF_10_ = 1.27, PD = 0.94, ROPE = 0.22), and PANSS general (ρ = 0.23, 95% CI [-0.03, 0.47], BF_10_ = 1.91, PD = 0.95, ROPE = 0.16), but not PANSS negative (ρ = -0.10, 95% CI [-0.35, 0.15], BF_10_ = 0.58, PD = 0.78, ROPE = 0.43). These findings suggest that individuals with initially lower total, positive, and general symptom severity were more likely to exhibit brain recovery. We did not find evidence of a relationship between brain age gap reduction and the baseline cognitive impairments measured by the composite score, TMT mean, DST working memory, VLMT short-term memory, or VLMT long-term memory (see Figure 6).

**Figure 6.**
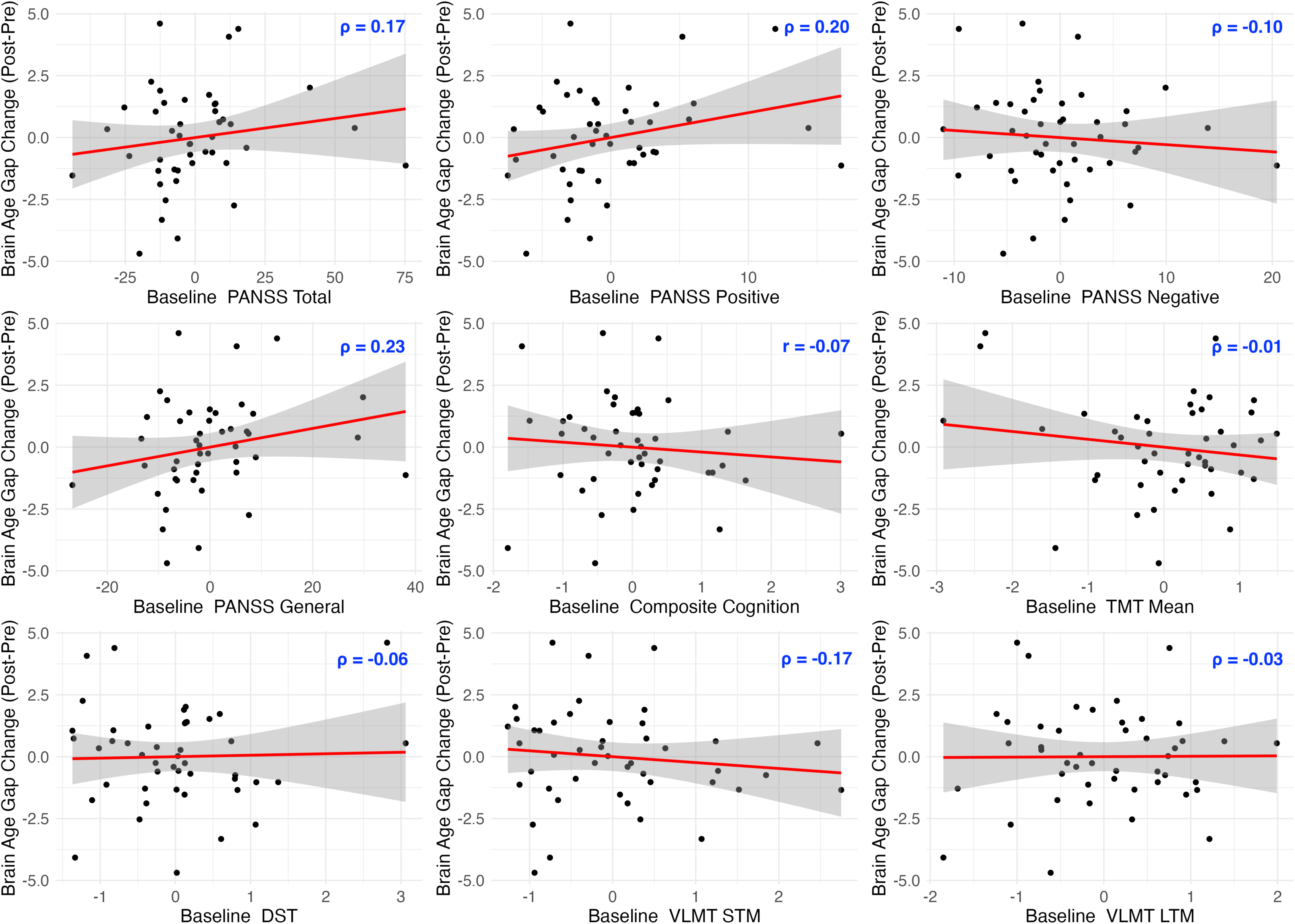
Partial Correlations: Clinical Values at Baseline vs. Brain Age Gap Change (Adjusted Values) *Note.* The graphs display the partial correlations between change in brain age gap (post-pre, larger values indicating accelerated brain aging) and baseline clinical (larger values indicating more severe symptoms) and cognitive variables (larger values reflecting better cognition). All variables are adjusted for covariates, and thus, the values shown represent residuals. STM: short-term memory; LTM: long-term memory.

Our results showed no influence of the schizophrenia polygenic risk score on brain age change.

### 3.3. Exploratory Findings

Exploring how the demographic variables relate to brain age gap change and brain age gap at baseline, we found strong evidence that patients with a higher BMI at baseline showed an increased brain recovery, denoted by a greater reduction in brain age gap (ρ = -0.36, 95% CI [- 0.57, -0.12], BF_10_ = 21.18, PD = 0.85, ROPE = 0.36). The baseline brain age gap, on the other hand, showed very strong evidence of a positive correlation with CPZ (ρ = 0.39, 95% CI [0.16, 0.61], BF_10_ = 34.21, PD = 1.00, ROPE = 0.01), whereby the brains of those patients who were receiving a higher medication dose were predicted to be older at baseline. For further exploratory results and the results on the Global Assessment of Functioning (GAF; Endicott et al., 1976), see supplementary materials.

## 4. Discussion

This study aimed to validate the brain age gap as a neurobiological outcome measure, a metric for post-exercise treatment response, and a predictor of clinical and cognitive improvements following physical exercise interventions in individuals with SSD.

We found no evidence for our first hypothesis, as the baseline brain age gap showed no association with aerobic fitness, clinical and cognitive parameters, or PRS. The inability to replicate previously reported relationships between fitness and the brain age gap likely stems from methodological heterogeneity in the literature. While some studies used BMI, waist circumference, heart rate, blood pressure, or cardiac stress test (Dunås et al., 2021; Wing et al., 2022), we used a lactate threshold test, which has its own limitations. For instance, blood lactate is an indirect, albeit more ethically permissible, measure of lactate formation in muscle tissue, which may not linearly map onto muscle levels (Armstrong and Welsman, 2007). Furthermore, its quantitative value, definition, and underlying mechanisms remain debated in the literature, despite being commonly regarded as a reliable indicator of cardiovascular fitness (Antonutto and Prampero, 1995; Boutcher, 1990; Faude et al., 2009).

Nonetheless, we found a positive correlation between BMI and brain age gap at baseline. This echoes the recent findings by Wing and colleagues (2024) showing that body fat percentage and visceral adipose tissue, but not fitness, predicted brain age gap change following a 6-month exercise intervention. Obesity is also a risk factor for increased brain aging in people with SSD (McWhinney et al., 2021, 2022) and in the healthy population (Beyer et al., 2019; Ronan et al., 2016) due to its disseminated impact on the brain (Chen et al., 2018; Kullmann et al., 2015), possibly mediated by adipocytokine- or fibrinogen-driven inflammation (Cazettes et al., 2011; Parimisetty et al., 2016; Wisse, 2004), cardiovascular strain (Cox et al., 2019; Goldstein et al., 2020; Jagust et al., 2005; Van Gaal et al., 2006), or lifestyle-related behaviors such as poor dietary habits, physical inactivity, or substance use (Kuo et al., 2013; Manu et al., 2015). In light of this, it is not surprising that patients with higher baseline BMI showed greater brain age improvement in our sample, likely due to their greater potential for exercise-induced enhancements. This observation parallels findings that structural and functional brain recovery follows interventions targeting obesity (Li et al., 2023). Future interventions in SSD can leverage these results to optimally recruit patients with comorbid obesity, who are most likely to benefit in terms of brain health. Promising approaches to simultaneously address obesity include integrated lifestyle modifications—focusing on exercise, diet (Agarwal et al., 2020; Aucoin et al., 2020), sleep (Ferrarelli, 2020; Gica and Selvi, 2021; Waite et al., 2020), and microbiome health (Dohnalová et al., 2022; Koblinsky et al., 2023), along with pharmacological options like semaglutide (Agarwal and Hahn, 2024). By prioritizing these strategies, treatment outcomes may be significantly enhanced.

The brain age gap at baseline did not relate to any clinical or cognitive parameters in our sample. Although a cross-sectional increase in brain age gap has been consistently reported in the literature, the exact link to symptom severity is equivocal (Constantinides et al., 2023; Demro et al., 2022; Haas et al., 2022; Hua et al., 2024; Kaufmann et al., 2019; Koutsouleris et al., 2014; Nenadić et al., 2017; Schnack et al., 2016; Shahab et al., 2019b; Tønnesen et al., 2020). Consistent with this, Haas et al. (2022) found no correlations between the brain age gap and symptom severity in early SSD. In contrast, McWhinney and colleagues (2021) reported a link between baseline brain age gap and negative symptom severity as well as functioning in FEP. One explanation could be the heterogeneity in the clinical presentation of SSD, such that this pattern is observed in subgroups but not in the overall SSD population. Indeed, Hua and colleagues (2024) found that experiential, but not expressive, negative symptom severity is associated with an increased brain age gap.

Another possibility is that increased brain age results from an early developmental trajectory of neurological impairments, termed early hit non-progressive, paralleling the neurodevelopmental hypothesis (Fatemi and Folsom, 2009; Marenco and Weinberger, 2000; Murray and Lewis, 1987b), with subsequent aging comparable to the healthy population. This model suggests that accelerated brain aging may be independent of current symptom severity, especially in post-acute samples like ours (Demro et al., 2022; Koutsouleris et al., 2014; Shahab et al., 2019c). Empirical support for this comes from an observational study, where FEP was characterized by an increased brain age gap at baseline but with analogous yearly brain aging rates compared to healthy controls (McWhinney et al., 2021). Contrary evidence also exists (Wang et al., 2021b), once again highlighting the heterogeneity, as many—but not all—patients fit into this model, underscoring the need for further research (Mjellem and Kringlen, 2001). In addition, a protective effect of antipsychotic agents against accelerated aging has been proposed, which can further complicate the relationship between symptom severity and brain age gap (Higgins-Chen et al., 2020; Schnack et al., 2016; Teeuw et al., 2021). For instance, severe symptoms treated with higher-dose medication may lead to a stabilization of brain aging. Alternatively, a history of severe symptoms could explain both the current larger brain age gap and higher medication dose, which is in turn effective in symptom reduction. Consistent with this, we observed a strong positive link between CPZ and the baseline brain age gap. Such a process may have obscured the relationship between the baseline brain age gap and clinical variables. A critical step forward in understanding these complex multivariate relationships and addressing the heterogeneity in the schizophrenia spectrum would be using stratification strategies, for instance, based on the progressiveness of the neural impairments, medical history, and medication dose.

Contrary to our hypothesis, we did not find an association between schizophrenia PRS and the baseline brain age gap. Only a few studies to date have examined this relationship. While Teeuw et al. (2021) reported a positive relationship in a mixed sample of 394 patients and controls, this finding did not survive Bonferroni corrections. Similarly, Constantinides et al. (2024) found null results in a sample of 189 participants from the general population. In contrast, our study focused exclusively on patients and incorporated a longitudinal exercise intervention, but the sample size (particularly in the longitudinal analysis testing our fifth hypothesis, linking PRS to brain age change, *n* = 21) limited our ability to detect a potential effect. Despite the much larger sample sizes, however, the studies mentioned above reported null results, prompting Constantinides and colleagues (2024) to suggest that structural MRI-derived brain age gap estimates may not be sensitive to schizophrenia PRS. Another plausible explanation, however, could be the low statistical power of these studies. Notably, with a much larger sample size of 18,088, Zhu et al. (2021) revealed a relationship between schizophrenia PRS and temporal and frontal, but not global, cortical volume. Accordingly, the direct effect of PRS on brain structure may be modest, requiring either very large sample sizes or region-focused local algorithms, as opposed to global ones like in this paper. In accord with this, a previous investigation of the Exercise2 sample found that PRS predicted post-exercise volume recovery in left hippocampal subfields of cornu ammonis4 and dentate gyrus (CA4/DG) but not in total hippocampus (Papiol et al., 2017). In a similar vein, the polygenic risk associated with specific cell types, oligodendrocyte precursor and radial glia cells, predicted less recovery in CA4/DG post-exercise in the same sample (Papiol et al., 2019). These findings suggest that the influence of genetic burden on brain recovery may be cell-type- or region-dependent.

Interestingly, Teeuw and colleagues (2021) reported an inverse relationship between schizophrenia PRS and epigenetic aging measured by DNA methylation, contrasting with their findings for the PRS and brain age gap. This suggests that different biological aging processes may diverge in their relationship with genetic risk factors. Taken together, these highlight the relevance of considering different algorithms as well as neuroimaging and biological modalities when assessing accelerated aging. Combining these approaches (e.g., epigenetic and brain aging, local and global brain aging) in future research may offer deeper insights into potentially independent mechanisms, enhancing both predictive accuracy and informativeness (Cole et al., 2019, 2018; Jansen et al., 2021; Rokicki et al., 2021; Teeuw et al., 2021).

We did not observe an omnibus decrease in the brain age gap after the exercise intervention. This contrasts with our hypothesis but parallels the results of a study on healthy older adults (Wing et al., 2024). This finding underscores the heterogeneity in individual treatment-response profiles, indicating that not everybody benefits from the same treatment. Future designs can incorporate comprehensive multi-domain interventions—addressing nutrition, microbiome, sleep, immunology, psychoeducation, social interactions, and mindfulness—to achieve higher response rates and synergistic effects on neuroplasticity (Abram et al., 2023; Chang et al., 2022; de Jong et al., 2019; Díaz-Zuluaga et al., 2017; Dohnalová et al., 2022; Firth et al., 2020, 2016, 2015; Lyman et al., 2014; Mrazek et al., 2016; Salzman et al., 2022; Sethi et al., 2024; Sivera et al., 2020; Solomon et al., 2021).

Our findings related to our third hypothesis showed that improvements, specifically in negative symptoms, were related to longitudinal reductions in brain age. This specificity may be attributed to several factors, including our sample characteristics and the distinct neurobiological and behavioral processes underlying different symptom clusters.

First, negative and positive symptoms are distinct clusters and are hypothesized to have different etiologies. A developmental frontal overpruning is argued to underlie negative and cognitive symptoms, while dopaminergic disruptions underpin positive symptoms (Howes and Shatalina, 2022). Conversely, the neurobiology underlying general psychopathology, as measured by the PANSS general, appears more diffuse, with changes in cortisol levels leading to broad metabolic effects, contributing to symptoms such as anxiety, somatic concern, and tension (Andrade et al., 2016; Iob and Steptoe, 2019). Moreover, on the behavioral level, negative symptoms specifically reflect dysfunctional avoidance, characterized by a lack of interest, social withdrawal, anhedonia (inability to experience pleasure), and avolition (severe lack of motivation). Improvement in this domain is particular as it represents a global lifestyle shift, manifesting in increased motivation and self-efficacy. While a sedentary lifestyle with minimal social engagement contributes to neural decline (Cai et al., 2023; Raichlen et al., 2023), exercise interventions promote an active, healthy, and social lifestyle, which can support brain recovery (Bradley et al., 2022; Chia et al., 2023; Liu et al., 2024). Alternatively, this effect could work in the opposite direction—or even bidirectionally—where neural recovery drives changes in behavior and lifestyle, reducing negative symptoms. Paralleling this argument, readiness or capability for lifestyle change may explain our findings concerning our fourth hypothesis: that lower baseline symptom severity predicts greater brain recovery, or conversely, that a lower brain age gap predicts improvement in composite and global cognition and long-term memory. Further evidence is required to make definitive claims, however, as the last two findings were only anecdotal. Our findings are nevertheless in accord with those of (Fan et al., 2024), which show that greater baseline symptom severity predicts an increase in the brain age gap, and that a higher baseline brain age gap predicts less functional recovery in response to risperidone treatment.

Second, we included post-acute patients in this study with at least two weeks of stable treatment on one or two antipsychotic agents, which are known to effectively reduce rates of major positive symptoms and general psychopathology (Haddad and Correll, 2018; Leucht et al., 2017). Consequently, patients tended to exhibit less severe positive symptoms compared to negative symptoms. The limited potential for further improvement in positive symptoms, largely addressed by antipsychotic treatment, may be less related to underlying brain structure due to the more episode-specific, dynamic, and transient nature of positive symptoms, often persisting for shorter durations than negative symptoms. In line with this argument, treatment response to antipsychotic medication correlates with advancements in the brain’s functional organization (Hadley et al., 2016; Li et al., 2020). In contrast, increasing severity of negative symptoms is associated with structural alterations, for instance, a longitudinal rise in cerebrospinal fluid volume and a volume reduction in frontal regions, left cerebellum, left posterior cingulate, and occipitoparietal cortices (Ho et al., 2003; McKechanie et al., 2016). Moreover, improvements in negative symptoms correlate not only with brain recovery assessed through functional imaging (Li et al., 2018), but also with the reversal of volume reductions in the left hippocampus, parahippocampus, and precuneus following brain stimulation (Hasan et al., 2017). Although longitudinal changes in positive symptoms have also been linked to structural changes in the brain, these often occur around disease onset and in concentrated areas like the perisylvian region or lateral ventricles. Conversely, negative-symptom-related morphological alterations are more widespread, spanning thalamocortical, temporal, and subcortical regions as well as the ventricles, occurring throughout the critical and chronic periods (Ho et al., 2003; Koutsouleris et al., 2008; Kurachi et al., 2018). However, caution is warranted in generalizing these results due to heterogeneity in study designs, sample characteristics and sizes, and the lack of targeted interventions (Dietsche et al., 2017; Mathalon et al., 2001). Our study uniquely contributes to the literature by drawing parallels between longitudinal structural neuroplasticity in response to a non-pharmaceutical intervention and distinctive symptom clusters. A fruitful realm of future investigation is to incorporate brain age algorithms utilizing functional MRI and EEG data, alongside structural MRI, to better capture dynamic treatment responses and connect functional neuroplasticity to improvements in specific symptom clusters.

As with negative symptom improvements, gains in composite cognition and long-term memory were also captured anecdotally in the brain age changes post-exercise. In line with this, cognitive decline related to aging and neurodegenerative disorders is linked to longitudinal brain atrophy (Carmichael et al., 2010; Jiang et al., 2016; Kramer et al., 2007; Mungas et al., 2005). In these studies, follow-up measurements were often taken at least 1 year after baseline, while we assessed changes after 3 to 6 months, which may explain the lower strength of evidence we observed. Indeed, in a short-duration cognitive training program lasting three to four weeks, stroke patients’ responses to the treatment did not reflect changes in the brain age gap (Richard et al., 2020). A 2-year cognitive enhancement therapy, on the other hand, reversed cognitive decline in early schizophrenia patients, where preservation of the left parahippocampal and fusiform gyrus and growth of the left amygdala predicted cognitive recovery (Eack et al., 2010). Shorter interventions also demonstrate this effect, with more pronounced changes observed in localized structural or functional domains rather than global structural ones. For instance, in a 3-month exercise intervention for healthy older adults, local grey matter plasticity in the middle frontal sulcus was related to enhanced cognitive functioning (Soshi et al., 2021). Similarly, a 4-month computerized cognitive training for schizophrenia patients led to increased functional efficiency in the middle frontal gyri, which correlated with working memory improvements (Subramaniam et al., 2014).

Taken together, these results highlight several critical factors: type of neuroimaging methods, diagnostic sample, duration and nature of the intervention, statistical power, and the specificity of the effects. Shorter interventions, such as ours, might primarily induce localized or functional changes, which precede global morphological changes (Isaac and Januel, 2016; Osmanlıoğlu et al., 2020). As our brain age predictions were based on global brain structure, we may have observed only low strength of evidence for these subtler local shifts. Despite our null findings for specific cognitive domains other than long-term memory, the negative correlation trends are descriptively in line with the previous findings. Evidence for the other cognitive domains could emerge with a larger sample size, a longer or multidomain intervention, or functional imaging techniques. Additionally, locally trained brain age models targeting specific regions of interest, such as the medial frontal cortex, may better capture subtle changes associated with cognitive improvements, specifically with working memory.

Long-term memory processes are dissociable from working memory (Grover et al., 2022; Simons and Spiers, 2003) and regulated by the medial temporal cortices and the hippocampus, regions known for their high plasticity (Cutsuridis and Yoshida, 2017). Notably, the hippocampus is one of the few areas in the adult brain where extensive neurogenesis is observed (Bruel- Jungerman et al., 2007). Furthermore, neuroplasticity in the hippocampal formation has been demonstrated in response to physical exercise interventions in individuals with SSD (Girdler et al., 2019; Maurus et al., 2019a; Pajonk et al., 2010; Roell et al., 2024; Woodward et al., 2018). The malleability of these regions and their pivotal role in long-term memory may explain our finding that, aside from composite cognition, long-term memory change was the only cognitive domain to correlate with brain age gap change. Further investigations are needed to establish a more robust link between improvements in specific cognitive domains and brain recovery, as our findings were anecdotal and existing research remains limited.

A limitation of our study is the inability to make causal claims about the effects of exercise due to the observational design of the study. Additionally, the add-on CACR in the Exercise2 sample hinders us from making any exercise-specific statements. Instead, we focused on examining the mechanistic processes underlying improvement in schizophrenia, using non-pharmaceutical interventional datasets—a context in which clinical recovery has been well-documented (Dauwan et al., 2016; Falkai et al., 2017; Firth et al., 2020, 2015; Haeger et al., 2019; Maurus et al., 2021, 2019a, 2019c; Roell et al., 2022; Schmitt et al., 2018). Future directions involve investigating the specificity of exercise, the interrelationship between positive health behaviors, and the directionality of the effects. On a related note, progressive brain aging in SSD could theoretically confound our longitudinal results—given the absence of a no-exercise control group. However, as previously mentioned, current evidence suggests that brain age in SSD is advanced but non-progressive (Demro et al., 2022; McWhinney et al., 2021). Nonetheless, some argue that progressive brain aging may occur, particularly in FEP (Dietsche et al., 2017; Lieberman et al., 2001). This emphasizes the need for further investigation, especially since our sample primarily did not include this population, making it less likely that our results are affected by such a process. Our claims are also limited by our method of choice for determining the biological age. The growing interest in bodily ages above and beyond the morphology and function of the brain calls for a more holistic approach, including blood indicators within the measures (Cole et al., 2019; Han et al., 2019; Teeuw et al., 2021). We acknowledge that age, sex, medication, education, BMI, and smoking can influence biological age indicators, but we were unable to statistically account for smoking due to missing data for most of our sample, possibly introducing some unexplained variability.

To our knowledge, this study is the first to show that the brain age gap, as a singular neurological parameter, is sensitive to aspects of treatment response in SSD. Our results suggest that brain age prediction could serve as a valuable tool for quantifying neuroplasticity in response to treatment in severe mental illness. Furthermore, our findings stress the prominence of the modifiable lifestyle factors in brain recovery. BMI and clinical variables, but not PRS, were related to brain age variables. This suggests that substantial opportunities for rehabilitation exist in severe mental illness, with low-cost, non-pharmaceutical, and add-on interventions holding significant potential to improve outcomes and enhance long-term recovery.

We identified BMI as a key variable influencing brain health, underscoring its potential role in future treatment strategies. Our findings also emphasize the importance of brain health in circumventing negative symptoms in post-acute SSD. These insights pave the way for developing stratification strategies based on biomarkers underlying comorbid obesity or severe negative symptoms, which are essential as personalized medicine gains traction. Such strategies can optimize patient allocation and enhance treatment outcomes, addressing the multifaceted and heterogeneous nature of SSD. Furthermore, we highlight promising future directions, including combined lifestyle interventions—such as diet, exercise, social interactions—and targeted pharmacological options like semaglutide. Ultimately, we provide evidence supporting the use of brain age prediction as a surrogate outcome metric for tracking clinical progress and brain recovery in clinical trials. Moving forward, we advocate for the integration of multimodal brain imaging and assessments of other biological ages to enhance our understanding and treatment of severe mental illness.

## Data Availability

Explicit permission for data sharing was not included in the informed consent obtained during the data collection phase. As a result, and due to the sensitive nature of the clinical data, we are unable to share the data publicly. However, the data can be made available to individual researchers upon request. Researchers interested in accessing the data may contact us directly to discuss potential arrangements.

## Acknowledgements

We express our appreciation to the Clinical Open Research Engine (CORE) at the University Hospital LMU (Munich, Germany) for providing the computational infrastructure to run the CPU-intensive MRI analysis pipelines.

## Author contributions

Conceptualization: IM, LR, DY; Data curation: DY, LR, SP, JHC, BM; Formal analysis: DY; Funding acquisition: PF, AS, DY; Methodology: LR, DY, JHC, DK; Project administration: PF, AS, IM, DK, LR, BM; Resources: LR, JHC; Software: DY, LR, JHC, Supervision: PF, AS, IM, LR; Visualization: DY; Writing – original draft: DY; Writing – review and editing: DY, LR, IM, DK, JHC, SP, AS, PF.

