## Supplementary Materials for "Brain Age Gap Reduction Following Physical Exercise Mirrors Negative Symptom Improvement in Schizophrenia Spectrum Disorders"

#### Supplementary Methods

Exclusion criteria were severe comorbidities, drug abuse, and pregnancy. Exercise2 had the additional inclusion criteria of age between 18 and 60 years, a history of at least 2 psychotic episodes, stable antipsychotic for 2 weeks before and during the whole study period, and verbal IQ > 85. ESPRIT, on the other hand, included patients aged 18 to 65 years, a total Positive and Negative Syndrome Scale (PANSS) score less than or equal to 75 (post-acute phase indicator), and stable treatment with one or two antipsychotics.

We administered three cognitive tests: Verbal Learning and Memory Test (VLMT) (Helmstaedter and Durwen, 1990), Digit Span Test (DST) (Tewes, 1994), and the Trail Making Test (TMT) A and B (Reitan, 1985).

##### *VLMT*

In this test, the examiner read a list of 15 words, and participants tried to recall as many as possible in any order. This process was repeated five times (VLMT-1st to VLMT-5th), and the total of correctly recalled words across these trials was calculated (VLMT-sum). After the fifth trial, an interference list of 15 new words was introduced, which participants also attempted to recall (VLMT-inter). Without repetition, participants were then asked to recall words from the first list (VLMT-6th). Following a 20-minute delay, in which other cognitive tests were administered, participants again attempted to recall words from the first list (VLMT-7th). The difference in recall between the fifth and seventh trials was calculated (VLMT-diff). Lastly, the examiner read 50 words, including words from the initial list and interference list, and participants indicated if each word belonged to the first list (VLMT-recog). VLMT-1 and VLMT-inter were Z-standardized, averaged, and re-standardized to create the VLMT short-term memory variable. At the same time,

VLMT-6 and VLMT-7 were similarly processed to yield the VLMT long-term memory variable. Higher scores indicate greater recall and, hence better memory.

***DST***

For DST-forward, the examiner read sequences of numbers with increasing length, and participants repeated them in the same order until they missed two sequences of the same length. In DST-backward, participants repeated the sequences in reverse order. For this study, we only utilized DST-backward, which was scored based on the number of correct trials and Z-standardized to indicate working memory performance. Higher scores indicate better performance.

***TMT***

In TMT-A, participants connected numbers from 1 to 25 sequentially as quickly and accurately as possible without lifting the pencil. In TMT-B, they alternated between numbers and letters, following the sequence until reaching the number 13 (e.g., 1-A-2-B, etc.). The time taken in seconds was measured. TMT-A and -B were standardized, averaged, re-standardized, and negated by sign inversion to create the TMT mean variable. Higher scores indicate better performance.

Finally, we computed the composite cognitive score by averaging the TMT mean, VLMT short-term memory, VLMT long-term memory, and standardized DST working memory scores, followed by standardizing this average.

***Imputation of Variables***

Our criteria led to the imputing of 1% of the data for CPZ and education years; %14 of BMI; %0.5 of PANSS total, positive, negative, and general; %2.5 of VLMT 1, VLMT 6, VLMT 7, VLMT int; %2 of TMT A; and 0.5 of TMT B.

**Supplementary Results**

***Patient Characteristics and Exploratory Findings***

Age and brain-predicted age were highly correlated ($\rho = 0.66$, 95% CI [0.56, 0.75], $BF_{10} > 100$, PD = 1.00, ROPE = 0.00, see Figure S.1).

Inspecting how the demographic variables relate to brain age gap change and brain age gap at baseline, we found strong evidence that patients with a higher BMI at baseline showed an increased brain recovery, denoted by a greater reduction in brain age gap ($\rho = -0.36$, 95% CI [-0.57, -0.12], $BF_{10} = 21.18$, PD = 0.85, ROPE = 0.36) (see Figure S.2). The baseline brain age gap, on the other hand, showed very strong evidence of a negative correlation with age ($\rho = -0.53$, 95% CI [-0.69, -0.33], $BF_{10} = 2903.48$, PD = 1.00, ROPE = 0.00) and a positive correlation with CPZ ($\rho = 0.39$, 95% CI [0.16, 0.61], $BF_{10} = 34.21$, PD = 1.00, ROPE = 0.01), indicating that the brains of those patients, who were younger and receiving a higher medication dose were predicted to be older by the algorithm at baseline (see Figure S.3). The other variables did not predict brain age gap at baseline or its change post-exercise (0 < all $BF_{10}$ 's < 1).

Figure S.4 shows the partial Spearman rank correlations between all main baseline variables.

**GAF Findings.** Baseline GAF and baseline brain age gap showed anecdotal evidence of a negative correlation ($\rho = -0.14$, 95% CI [-0.30, 0.21], $BF_{10} = 1.14$, PD = 0.96, ROPE = 0.31; see Figure S.5.A). Improvement in GAF and change in brain age gap did not correlate ($r = -0.11$, 95% CI [-0.35, 0.16], $BF_{10} = 0.59$, PD = 0.80, ROPE = 0.39; see Figure S.5.B). The baseline brain age gap did not predict improvement in GAF ($r = 0.09$, 95% CI [-0.19, 0.34], $BF_{10} = 0.54$, PD = 0.76, ROPE = 0.43; see Figure S.5.C). Baseline GAF did not predict improvement in brain age gap ($\rho = -0.10$, 95% CI [-0.35, 0.16], $BF_{10} = 0.56$, PD = 0.78, ROPE = 0.46; see Figure S.5.D). GAF scores increased post-exercise ($M_{pre}$ = 62.80, $SD_{pre}$ = 11.20; $M_{post}$ = 70.40, $SD_{post}$ = 10.10; $BF_{10}$ = 6696.43).

***Association between Baseline Brain Age Gap, Aerobic Fitness, and Clinical Measures***

A Bayesian partial Spearman rank correlation revealed no link at baseline between aerobic fitness and brain age gap (ρ = 0.07, 95% CI [-0.11, 0.26], $BF_{10}$ = 0.39, PD = 0.76, ROPE = 0.61, see Figure S.6).

Brain age gap at baseline was also not correlated with any of the clinical parameters, namely PANSS Total (ρ = 0.05, 95% CI [-0.11, 0.22], $BF_{10}$ = 0.31, PD = 0.71, ROPE = 0.69), PANSS Positive (ρ = 0.03, 95% CI [-0.14, 0.19], $BF_{10}$ = 0.28, PD = 0.65, ROPE = 0.73), PANSS Negative (ρ = 0.05, 95% CI [-0.10, 0.22], $BF_{10}$ = 0.32, PD = 0.72, ROPE = 0.70), or PANSS General (ρ = 0.06, 95% CI [-0.12, 0.21], $BF_{10}$ = 0.32, PD = 0.73, ROPE = 0.67) as tested by Bayesian partial Spearman correlations (see Figure S.7).

At baseline, Pearson partial correlations revealed no significant relationship between the brain age gap and composite cognitive scores ($r$ = 0.06, 95% CI [-0.12, 0.21], $BF_{10}$ = 0.33, PD = 0.75, ROPE = 0.68). Neither did the partial Spearman correlations show a relationship between BrainAGE at baseline and individual cognitive domains TMT mean (ρ = -0.03, 95% CI [-0.19, 0.13], $BF_{10}$ = 0.28, PD = 0.64, ROPE = 0.74), DST.wm (ρ = 0.06, 95% CI [-0.10, 0.22], $BF_{10}$ = 0.36, PD = 0.78, ROPE = 0.63), VLMT short term memory (ρ = -0.01, 95% CI [-0.18, 0.15], $BF_{10}$ = 0.26, PD = 0.54, ROPE = 0.77), and VLMT long term memory (ρ = 0.13, 95% CI [-0.03, 0.28], $BF_{10}$ = 0.85, PD = 0.94, ROPE = 0.38). These results indicated that the brain age gap at baseline was not related to global cognition, working memory, verbal short-term memory, or verbal long-term memory, respectively (see Figure S.8).

Brain age gap scores at baseline and Polygenic Risk Scores were also not correlated ($r$ = 0.07, 95% CI [-0.17, 0.27], $BF_{10}$ = 0.41, PD = 0.70, ROPE = 0.55; see Figure S.9).

***Longitudinal Changes Following Physical Exercise Intervention***

To examine the longitudinal changes in the brain age gap in the context of physical exercise, we ran a Bayesian linear mixed-effects model, which did not provide evidence suggesting an effect of the session, demonstrating no overall change in the brain age gap post-exercise with time ($\beta$ = 0.13, 95% CI [-0.44, 0.71], $BF_{10}$ = 0.31). Similarly, there was no effect of the number of attended trainings ($\beta$ = -0.03, 95% CI [-0.18, 0.12], $BF_{10}$ = 0.81) or chlorpromazine-equivalent dose ($\beta$ = 0.00, 95% CI [0.00, 0.01], $BF_{10}$ = 0.02), or education years ($\beta$ = -0.22, 95% CI [-0.72, 0.29], $BF_{10}$ = 0.37) on the brain age gap. On the other hand, we found very strong evidence of the predictor age ($\beta$ = -0.48, 95% CI [-0.67, -0.30], $BF_{10} > 1000$) and BMI ($\beta$ = 0.26, 95% CI [0.12, 0.39], $BF_{10}$ = 134.81) and anecdotal evidence of sex ($\beta$ = 0.95, 95% CI [-0.90, 2.73], $BF_{10}$ = 1.71), study ($\beta$ = -1.35, 95% CI [-3.22, 0.53], $BF_{10}$ = 2.44), and group ($\beta$ = -0.61, 95% CI [-2.34, 1.18], $BF_{10}$ = 1.16). These results indicate that an increase in age, but a decrease in BMI, was related to a lower brain age gap. Participants from the Exercise2 study, males, and people assigned to the AET group had a higher brain age gap (see Figures S.10 and S.11).

A partial Spearman rank correlation revealed no evidence that brain age gap change and PANSS total change are related ($\rho$ = 0.11, 95% CI [-0.15, 0.36], $BF_{10}$ = 0.59, PD = 0.80, ROPE = 0.42). PANSS positive change ($\rho$ = -0.13, 95% CI [-0.38, 0.12], $BF_{10}$ = 0.71, PD = 0.84, ROPE = 0.37) and PANSS general change ($\rho$ = 0.06, 95% CI [-0.20, 0.32], $BF_{10}$ = 0.46, PD = 0.66, ROPE = 0.50) were also not correlated with brain age gap change. On the other hand, a partial Pearson correlation demonstrated moderate evidence that PANSS negative change ($r$ = 0.27, 95% CI [0.02,

0.50], $BF_{10}$ = 3.22, PD = 0.98, ROPE = 0.10) predicted brain age gap change, pointing to the link between post-exercise clinical improvement and brain recovery.

We found anecdotal evidence of a negative relationship between brain age gap change and a change in composite cognitive score (ρ = -0.26, 95% CI [-0.50, -0.01], $BF_{10}$ = 2.60, PD = 0.97, ROPE = 0.11) and VLMT long-term memory ($r$ = -0.25, 95% CI [-0.49, -0.01], $BF_{10}$ = 2.36, PD = 0.97, ROPE = 0.12). Hence, brain recovery post-exercise was related to improvements in general cognition and long-term memory. There was no evidence that brain age gap change is associated with a change in TMT mean (ρ = -0.06, 95% CI [-0.31, 0.19], $BF_{10}$ = 0.48, PD = 0.69, ROPE = 0.49), DST working memory (ρ = -0.12, 95% CI [-0.36, 0.14], $BF_{10}$ = 0.62, PD = 0.81, ROPE = 0.40), VLMT short-term memory ($r$ = -0.13, 95% CI [-0.39, 0.12], $BF_{10}$ = 0.66, PD = 0.83, ROPE = 0.38).

We inspected the predictors of treatment response and found that brain age gap at baseline was not related to the subsequent change in PANSS total (ρ = -0.02, 95% CI [-0.29, 0.23], $BF_{10}$ = 0.43, PD = 0.55, ROPE = 0.54), PANSS positive (ρ = 0.11, 95% CI [-0.13, 0.37], $BF_{10}$ = 0.61, PD = 0.80, ROPE = 0.41), PANSS negative ($r$ = -0.15, 95% CI [-0.40, 0.10], $BF_{10}$ = 0.78, PD = 0.86, ROPE = 0.32), or PANSS general (ρ = 0.04, 95% CI [-0.23, 0.28], $BF_{10}$ = 0.44, PD = 0.60, ROPE = 0.53). Examining the cognitive domain revealed anecdotal evidence of lower brain age gap at baseline to predict greater improvement in composite cognitive score (ρ = -0.22, 95% CI [-0.47, 0.02], $BF_{10}$ = 1.71, PD = 0.95, ROPE = 0.17), TMT mean change (ρ = -0.23, 95% CI [-0.47, 0.02], $BF_{10}$ = 1.80, PD = 0.96, ROPE = 0.15), and VLMT long-term memory change ($r$ = -0.21, 95% CI [-0.44, 0.06], $BF_{10}$ = 1.39, PD = 0.94, ROPE = 0.20). However, no evidence was found for DST working memory change (ρ = 0.06, 95% CI [-0.22, 0.31], $BF_{10}$ = 0.46, PD = 0.66, ROPE = 0.49) or VLMT short-term memory change ($r$ = -0.09, 95% CI [-0.33, 0.17], $BF_{10}$ = 0.54, PD = 0.76, ROPE = 0.45).

The relationship in the opposite direction concerning the correlation of baseline clinical characteristics to subsequent brain age change held anecdotally for baseline PANSS total (ρ = 0.17, 95% CI [-0.08, 0.42], $BF_{10}$ = 1.01, PD = 0.91, ROPE = 0.26), PANSS positive (ρ = 0.20, 95% CI [-0.07, 0.43], $BF_{10}$ = 1.27, PD = 0.94, ROPE = 0.22), and PANSS general (ρ = 0.23, 95% CI [-0.03, 0.47], $BF_{10}$ = 1.91, PD = 0.95, ROPE = 0.16), but not PANSS negative (ρ = -0.10, 95% CI [-0.35, 0.15], $BF_{10}$ = 0.58, PD = 0.78, ROPE = 0.43), indicating that those with initially lower total, positive, and general symptom severity were more inclined to show brain recovery. We did not find evidence of a relationship between brain age change and the baseline cognitive impairments measured by the composite score ($r$ = -0.07, 95% CI [-0.32, 0.20], $BF_{10}$ = 0.50, PD = 0.71, ROPE = 0.47), TMT mean (ρ = -0.01, 95% CI [-0.28, 0.24], $BF_{10}$ = 0.43, PD = 0.54, ROPE = 0.54), DST working memory (ρ = -0.06, 95% CI [-0.32, 0.21], $BF_{10}$ = 0.46, PD = 0.66, ROPE = 0.50), VLMT short-term memory (ρ = -0.17, 95% CI [-0.42, 0.09], $BF_{10}$ = 0.91, PD = 0.88, ROPE = 0.29), and VLMT long-term memory (ρ = -0.03, 95% CI [-0.30, 0.21], $BF_{10}$ = 0.44, PD = 0.59, ROPE = 0.52).

There was no link between polygenic risk scores and brain age change ($r$ = 0.14, 95% CI [-0.25, 0.48], $BF_{10}$ = 0.77, PD = 0.78, ROPE = 0.31; see Figure S.12).

### Supplementary Figures

Figure S.1

*Scatter Plot of Age vs. Brain-Predicted Age*

*Note.* The dashed line represents the identity line (y = x), and the red line represents the regression line.

Figure S.2

*Partial Spearman Rank Correlation Matrix of Brain Age Gap Change and Covariables*

*Note.* brain_age_gap_change_numeric: difference in brain age gap (post-exercise - baseline); education_years: total years of formal education; trainnum: number of training sessions attended; **: $p < 0.01$.

Figure S.3

*Partial Spearman Rank Correlation Matrix of Baseline Brain Age Gap and Covariables*

*Note.* education_years: total years of formal education; trainnum: number of training sessions attended; ***: $p < 0.001$.

Figure S.4

*Partial Spearman Rank Correlation Matrix of Baseline Variables*

*Note.* Correlation coefficients are presented in scientific notation where, for example, 8.77e−03 means $8.77 \times 10^{-3}$ or 0.00877. aerobic.fitness.2mmol: aerobic fitness operationalized as the exercise intensity at which blood lactate concentration reaches 2 mmol/l; education_years: total years of formal education; cogscore: composite cognitive score; *: $p < 0.05$; **: $p < 0.01$;
***: $p < 0.001$.

Figure S.5.A

*Partial Correlation Between Adjusted Baseline GAF and Baseline Brain Age Gap*

Figure S.5.B

*Partial Correlation Between Adjusted GAF Change and Brain Age Gap Change*

Figure S.5.C

*Partial Correlation Between Adjusted GAF Change and Baseline Brain Age Gap*

Figure S.5.D

*Partial Correlation Between Adjusted Baseline GAF and Brain Age Gap Change*

Figure S.6

*Partial Correlation Between Adjusted Baseline Aerobic Fitness and Baseline Brain Age Gap*

Figure S.7

*Partial Correlation Plots: Baseline Brain Age Gap vs. Baseline PANSS Scores*

*Note.* panss.pos: PANSS positive; panss.neg: PANSS negative; panss.gen: PANSS.general.

Figure S.8

*Partial Correlation Plots: Baseline Brain Age Gap vs. Baseline Cognitive Domains*

*Note.* tmt.mean: Trail Making Task Mean; dst.wm: Digit Span Test Working Memory; vlmt: Verbal Learning and Memory Test; stm: Short Term Memory, ltm: Long Term Memory.

Figure S.9

*Partial Correlation Between Adjusted Polygenic Risk Scores and Baseline Brain Age Gap*

Figure S.10

*Session Effects on Brain Age Gap by Study*

*Note.* Boxplots represent the brain age gap at baseline and post-exercise sessions (BL: baseline; 3m: 3-month post-exercise; 6m: 6-month post-exercise). Individual participants are depicted as dots, with lines connecting them to illustrate change. Participants showing brain recovery (a decreased brain age gap) are connected to their post-exercise session in green, while those with an increased brain age gap are shown in red. The left side displays the data from the Exercise2 and the right side from ESPRIT datasets.

Figure S.11

*Session Effects on Brain Age Gap by Training Group*

*Note.* The upper graph shows brain age gaps in the pooled data pre- and post-exercise grouped by the exercise group AET (Aerobic Endurance Training) on the left side and FSBT (Flexibility-Strength-Balance Training) on the right. The lower graph displays the same information by study, Exercise2 on the left and ESPRIT on the right. Dots represent individuals.

Figure S.12

*Partial Correlation: Polygenic Risk Score vs. Post-Exercise Brain Age Gap Change*

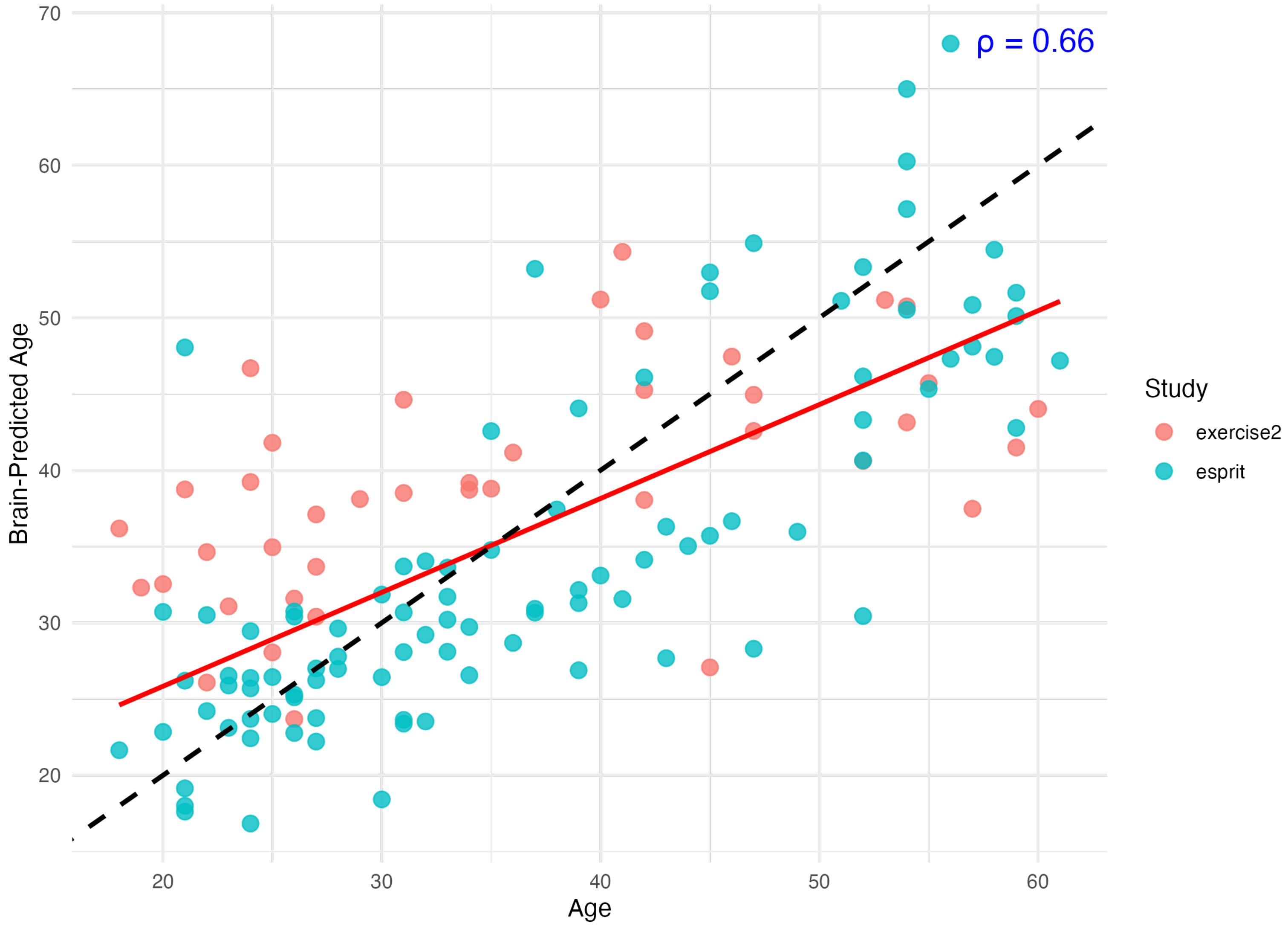

ρ = 0.66
Brain-Predicted Age
Age
Study
exercise2
esprit
20
30
40
50
60
70

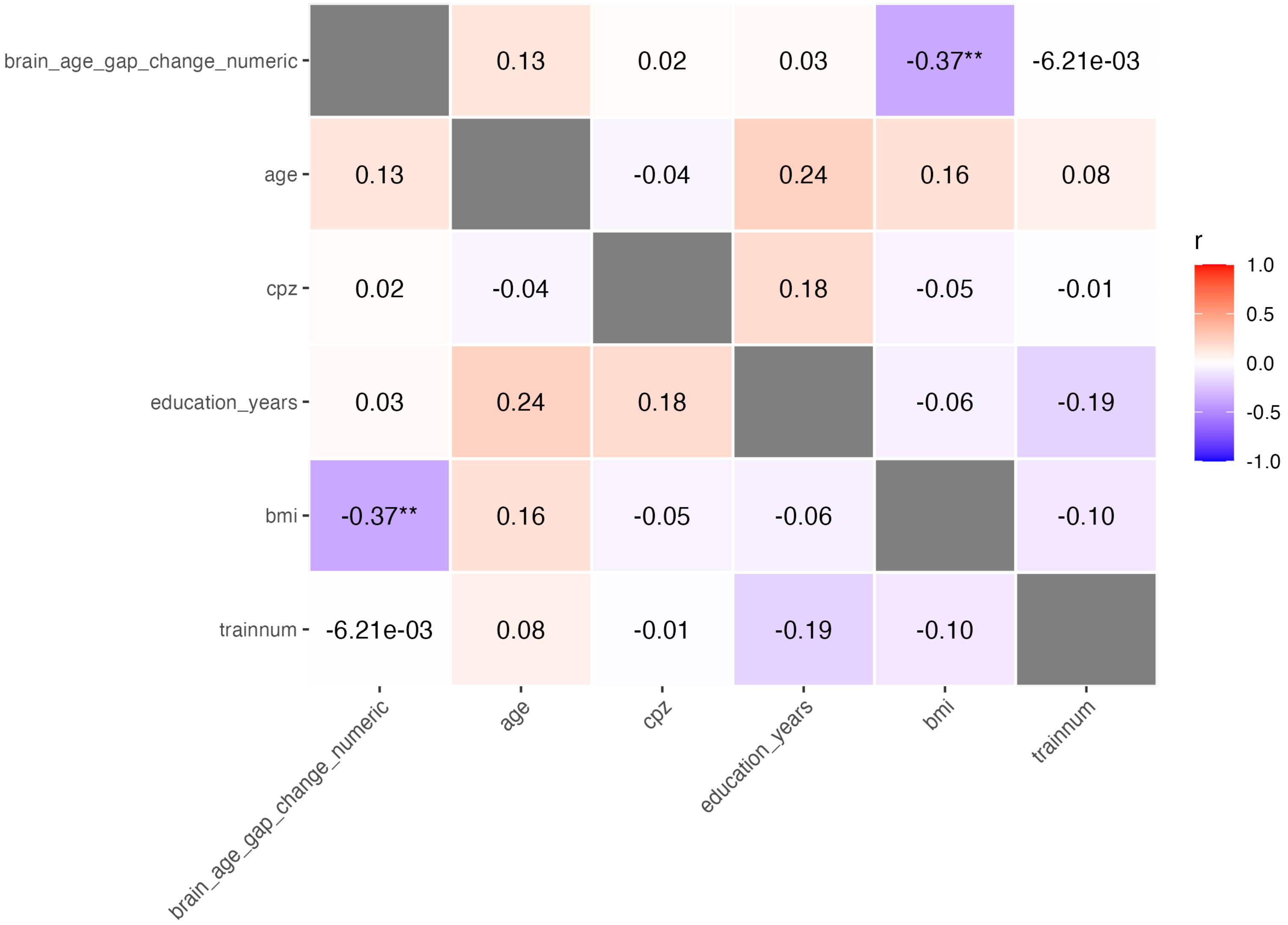
brain_age_gap_change_numeric
age
cpz
education_years
bmi
trainnum
0.13
0.02
0.03
-0.37**
-6.21e-03
0.13
-0.04
0.24
0.16
0.08
0.02
-0.04
0.18
-0.05
-0.01
0.03
0.24
0.18
-0.06
-0.19
-0.37**
0.16
-0.05
-0.06
-0.10
-6.21e-03
0.08
-0.01
-0.19
-0.10
r
1.0
0.5
0.0
-0.5
-1.0

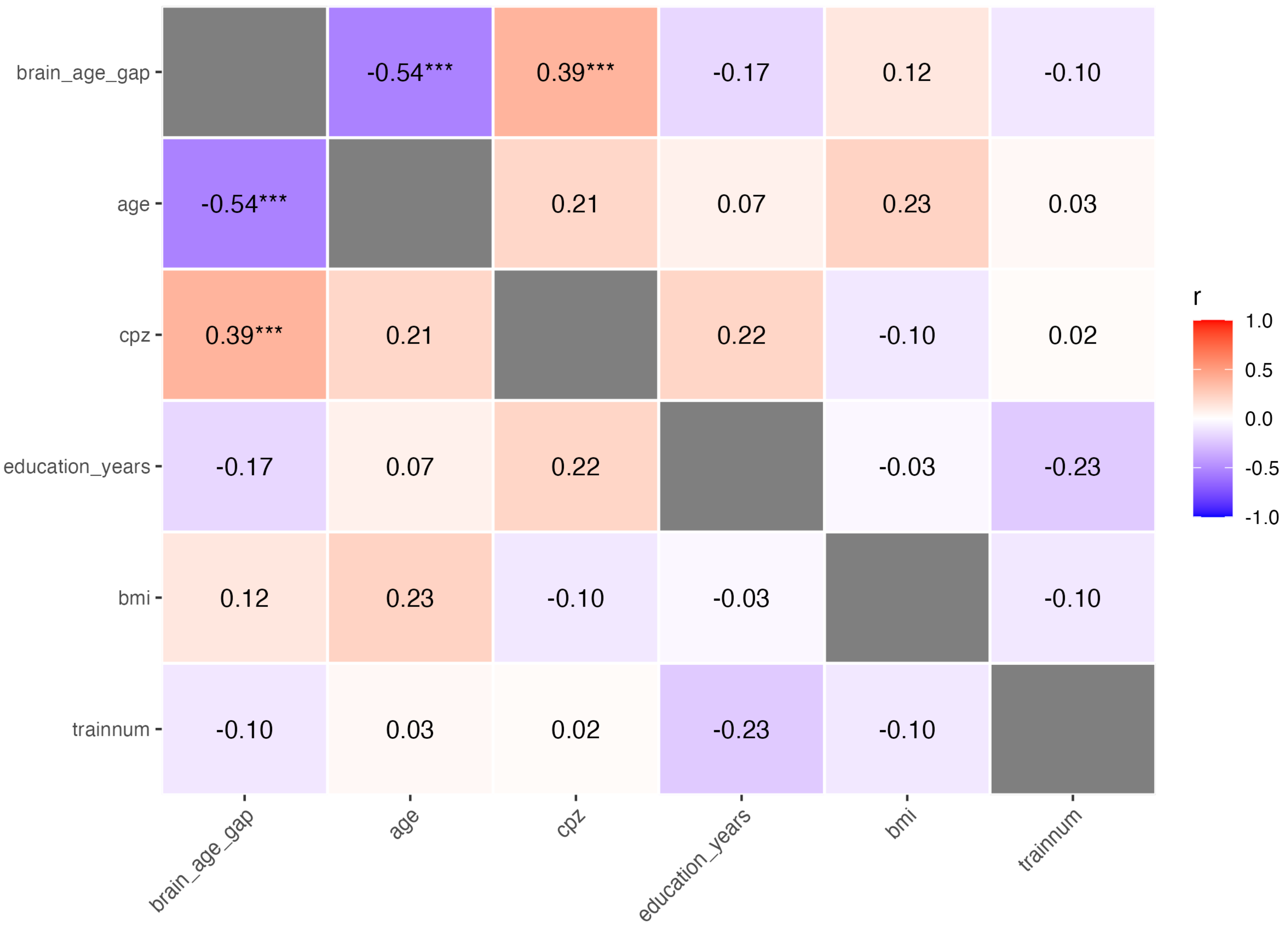

brain_age_gap
age
cpz
education_years
bmi
trainnum
-0.54***
0.39***
-0.17
0.12
-0.10
-0.54***
0.21
0.07
0.23
0.03
0.39***
0.21
0.22
-0.10
0.02
-0.17
0.07
0.22
-0.03
-0.23
0.12
0.23
-0.10
-0.03
-0.10
-0.10
0.03
0.02
-0.23
-0.10
r
1.0
0.5
0.0
-0.5
-1.0

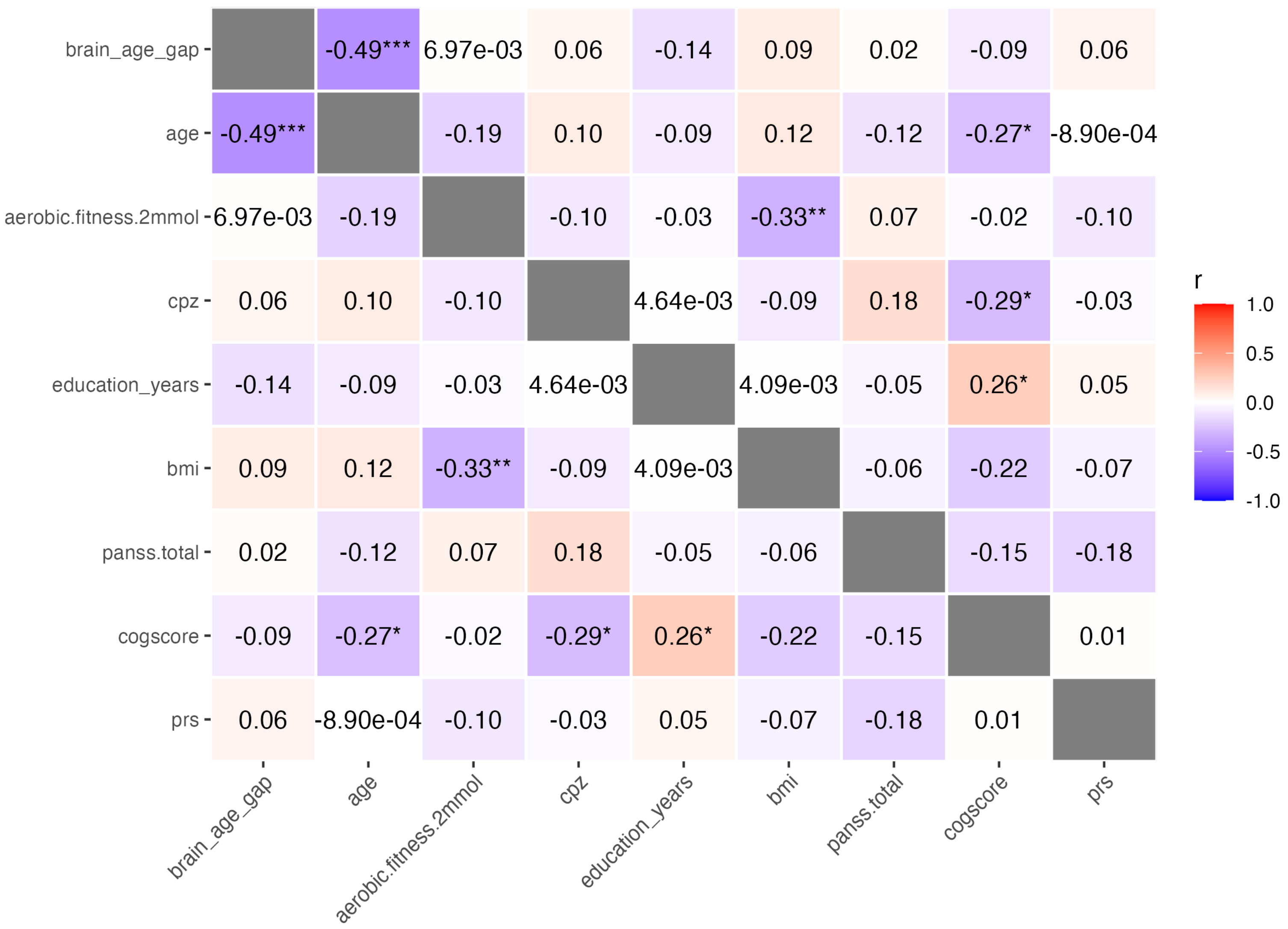

brain_age_gap
age
aerobic.fitness.2mmol
cpz
education_years
bmi
panss.total
cogscore
prs
-0.49***
6.97e-03
0.06
-0.14
0.09
0.02
-0.09
-0.19
0.10
0.12
-0.12
-0.27*
-8.90e-04
-0.10
-0.03
-0.33**
0.07
-0.02
4.64e-03
0.18
-0.29*
-0.05
0.26*
0.05
4.09e-03
-0.06
-0.22
-0.07
-0.15
-0.18
0.01
r
1.0
0.5
0.0
-0.5
-1.0

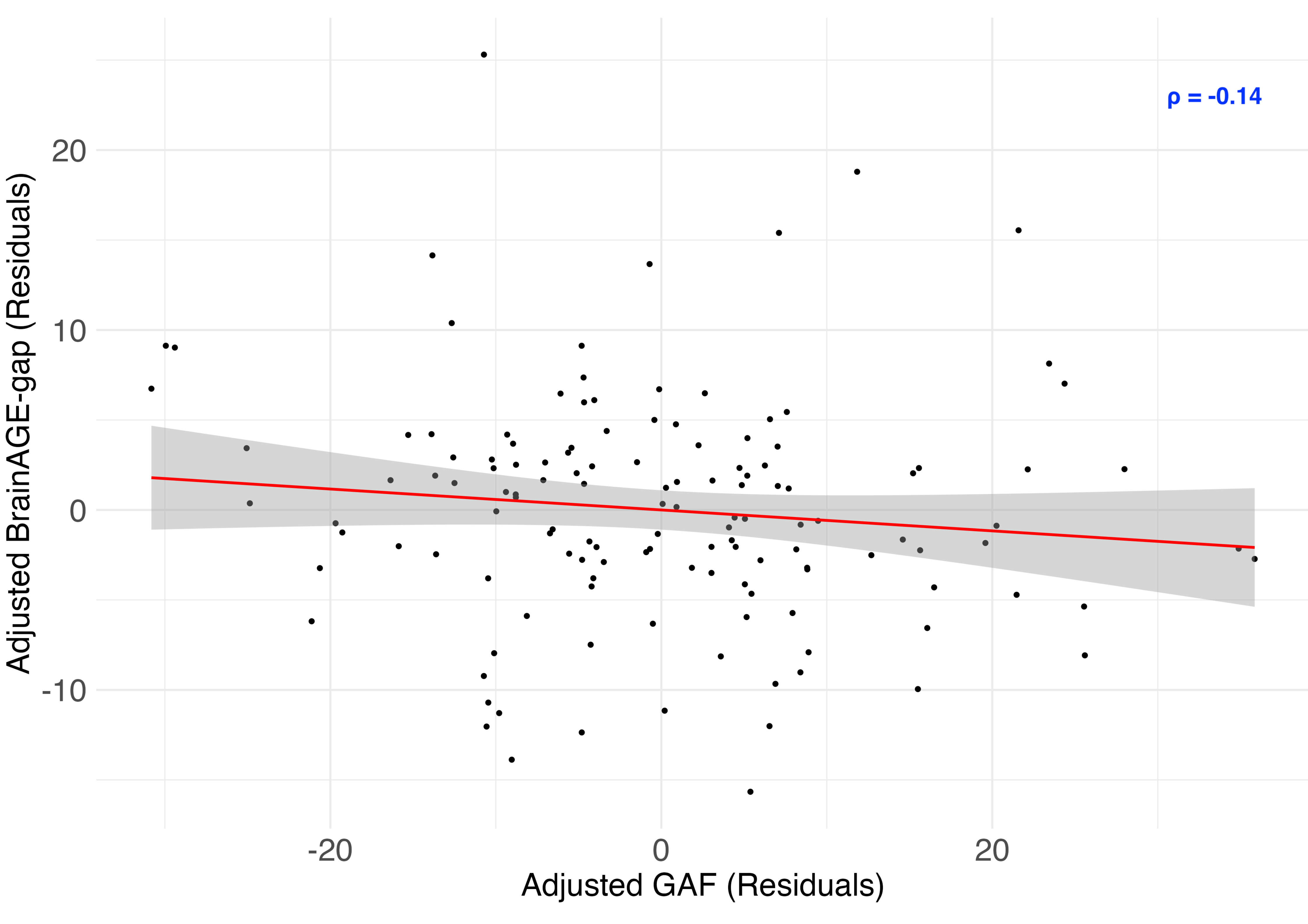
ρ = -0.14
Adjusted BrainAGE-gap (Residuals)
20
10
0
-10
-20
0
20
Adjusted GAF (Residuals)

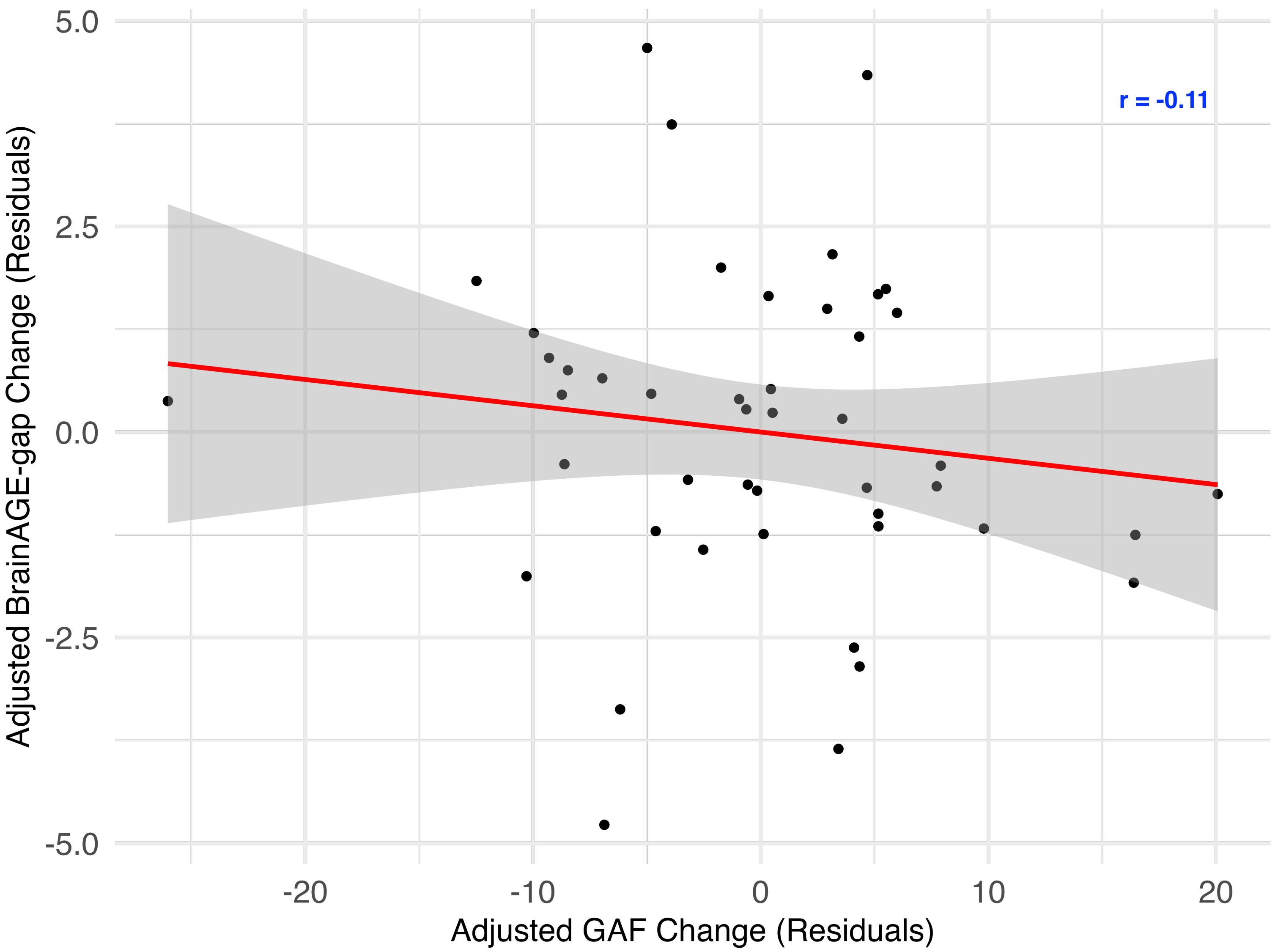

r = -0.11
5.0
2.5
0.0
-2.5
-5.0
-20
-10
0
10
20
Adjusted BrainAGE-gap Change (Residuals)
Adjusted GAF Change (Residuals)

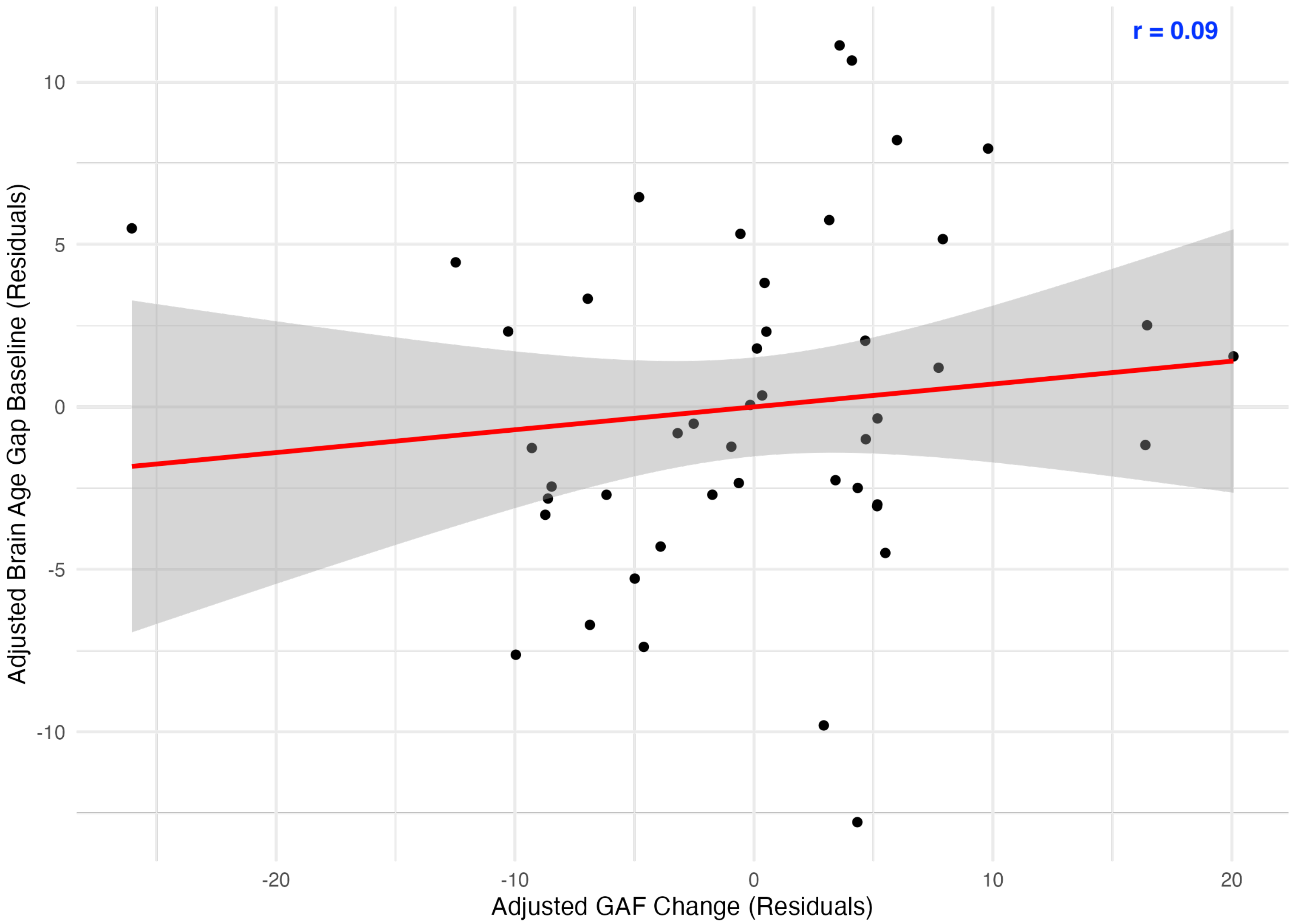

r = 0.09
10
5
0
-5
-10
Adjusted Brain Age Gap Baseline (Residuals)
-20
-10
0
10
20
Adjusted GAF Change (Residuals)

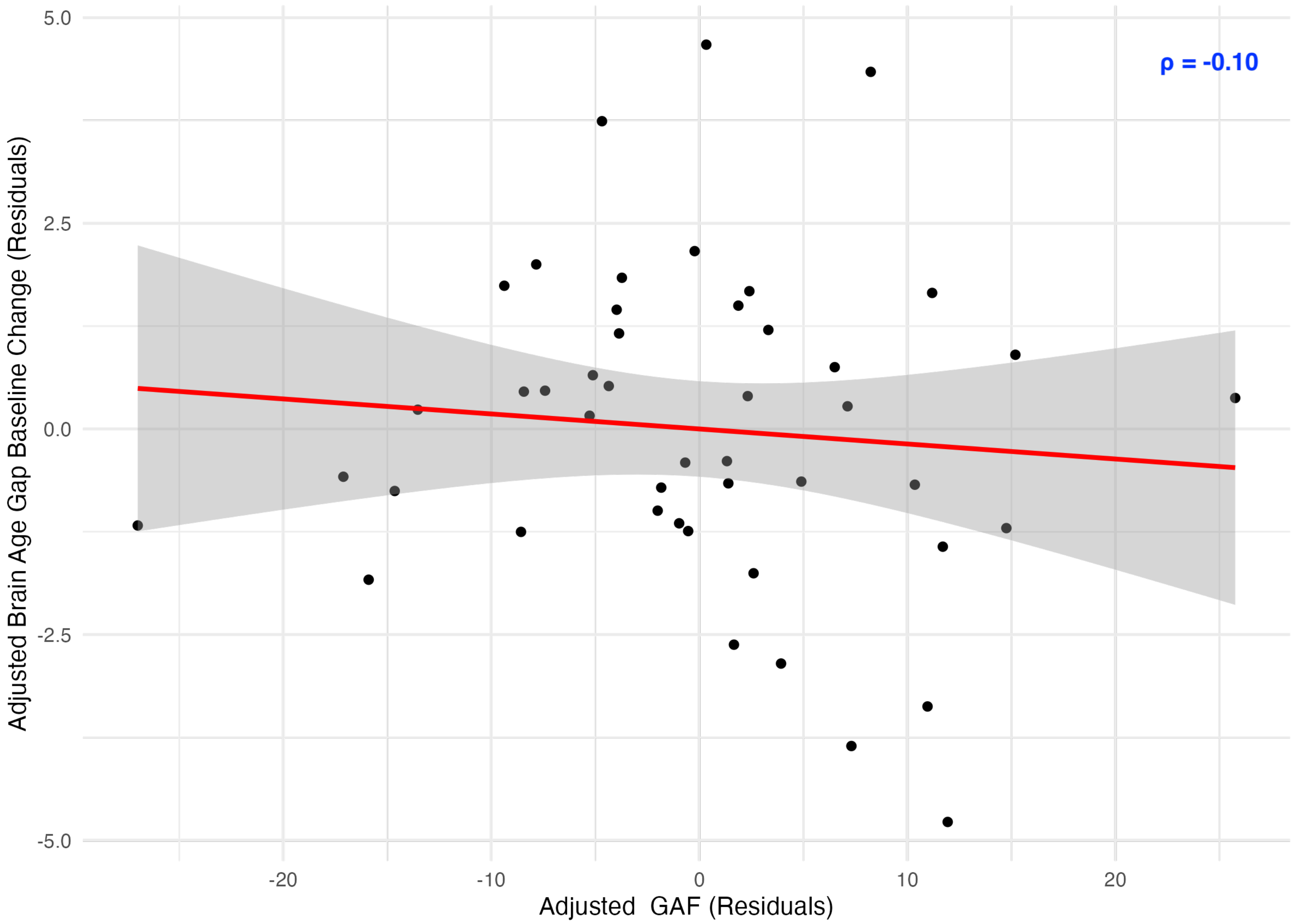

ρ = -0.10
5.0
2.5
0.0
-2.5
-5.0
-20
-10
0
10
20
Adjusted GAF (Residuals)
Adjusted Brain Age Gap Baseline Change (Residuals)

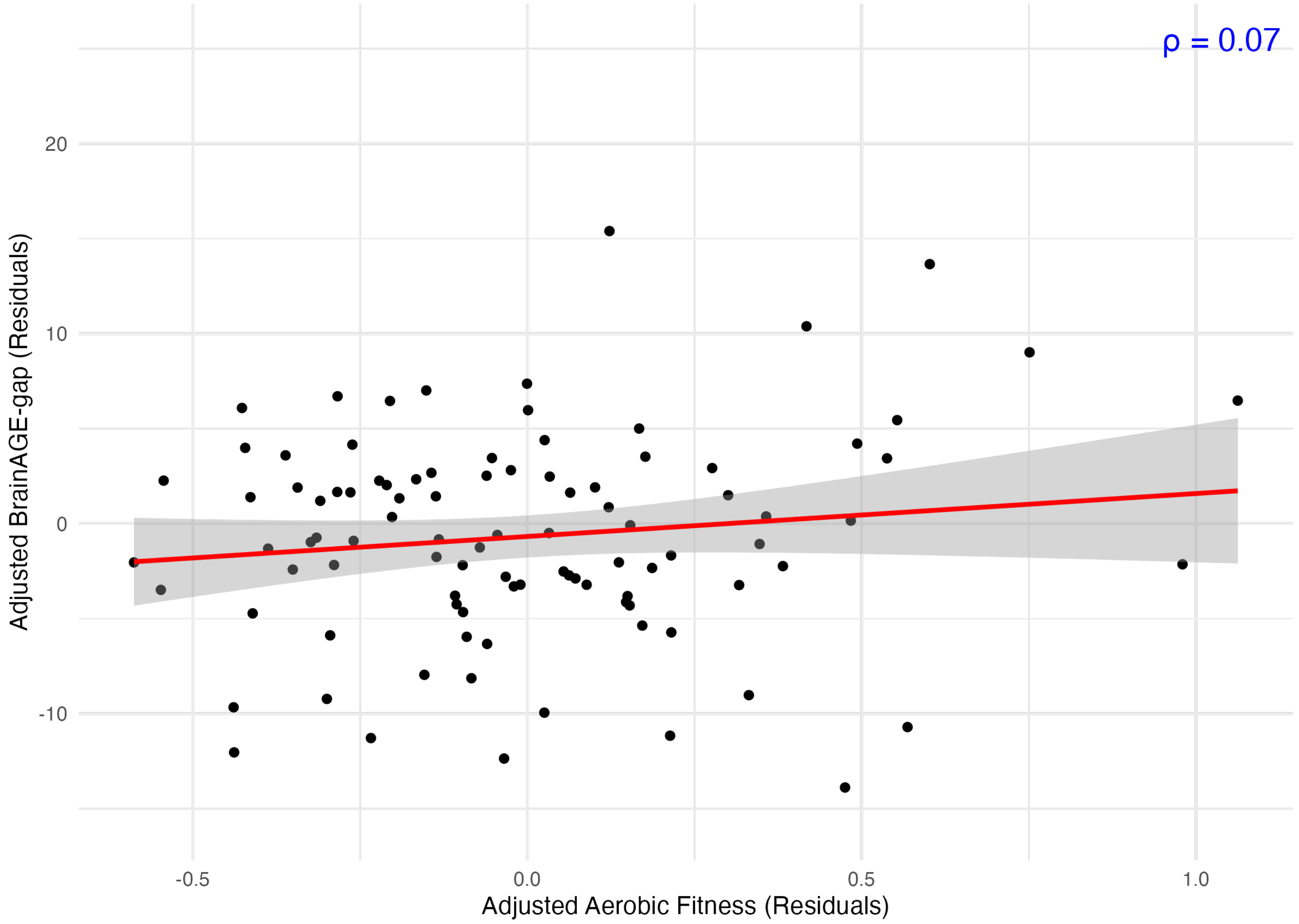

ρ = 0.07
20
10
0
-10
-0.5
0.0
0.5
1.0
Adjusted BrainAGE-gap (Residuals)
Adjusted Aerobic Fitness (Residuals)

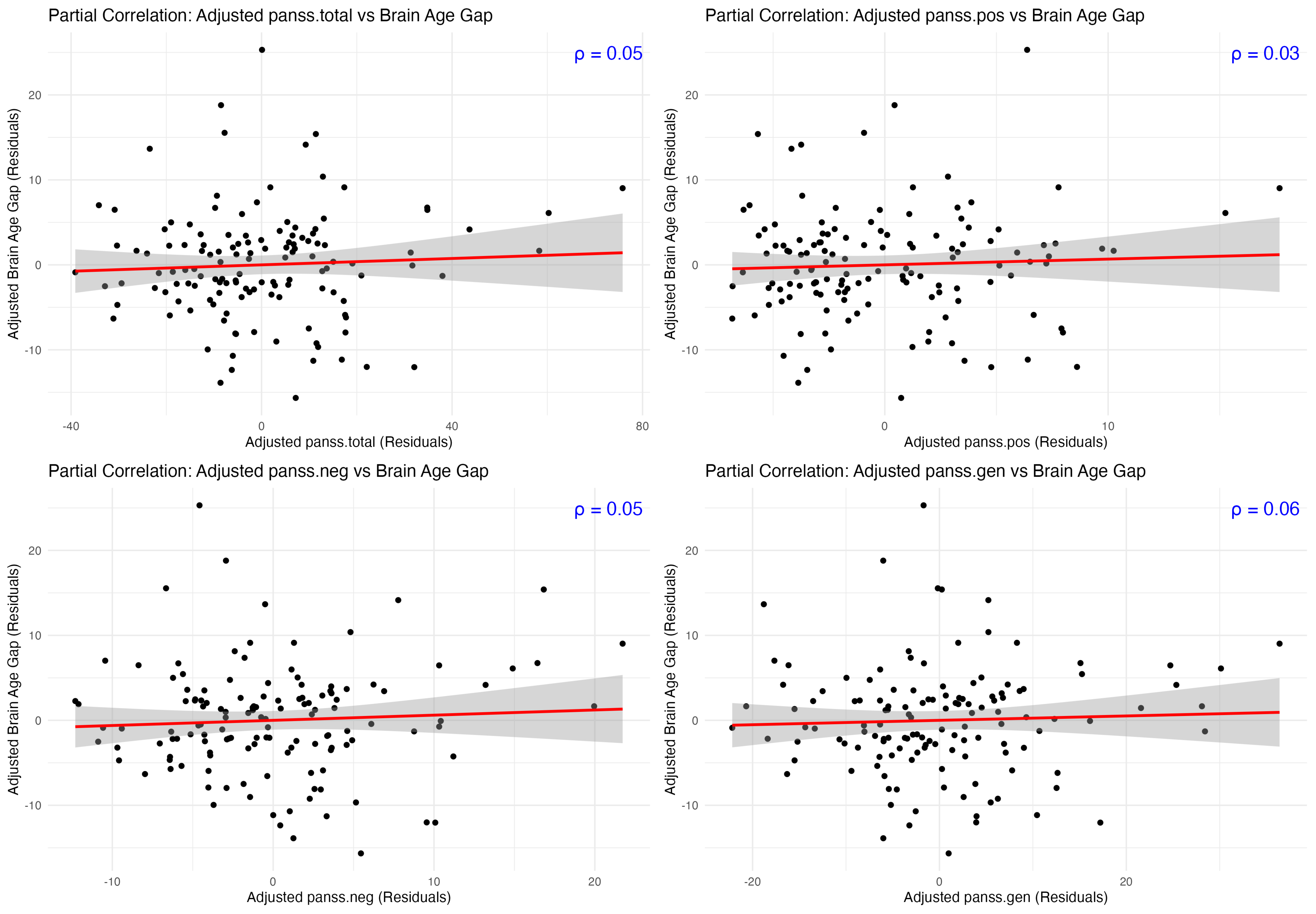

Partial Correlation: Adjusted panss.total vs Brain Age Gap
ρ = 0.05
Adjusted Brain Age Gap (Residuals)
Adjusted panss.total (Residuals)
-40
0
40
80
-10
0
10
20
Partial Correlation: Adjusted panss.pos vs Brain Age Gap
ρ = 0.03
Adjusted Brain Age Gap (Residuals)
Adjusted panss.pos (Residuals)
0
10
Partial Correlation: Adjusted panss.neg vs Brain Age Gap
ρ = 0.05
Adjusted Brain Age Gap (Residuals)
Adjusted panss.neg (Residuals)
-10
0
10
20
Partial Correlation: Adjusted panss.gen vs Brain Age Gap
ρ = 0.06
Adjusted Brain Age Gap (Residuals)
Adjusted panss.gen (Residuals)
-20
0
20

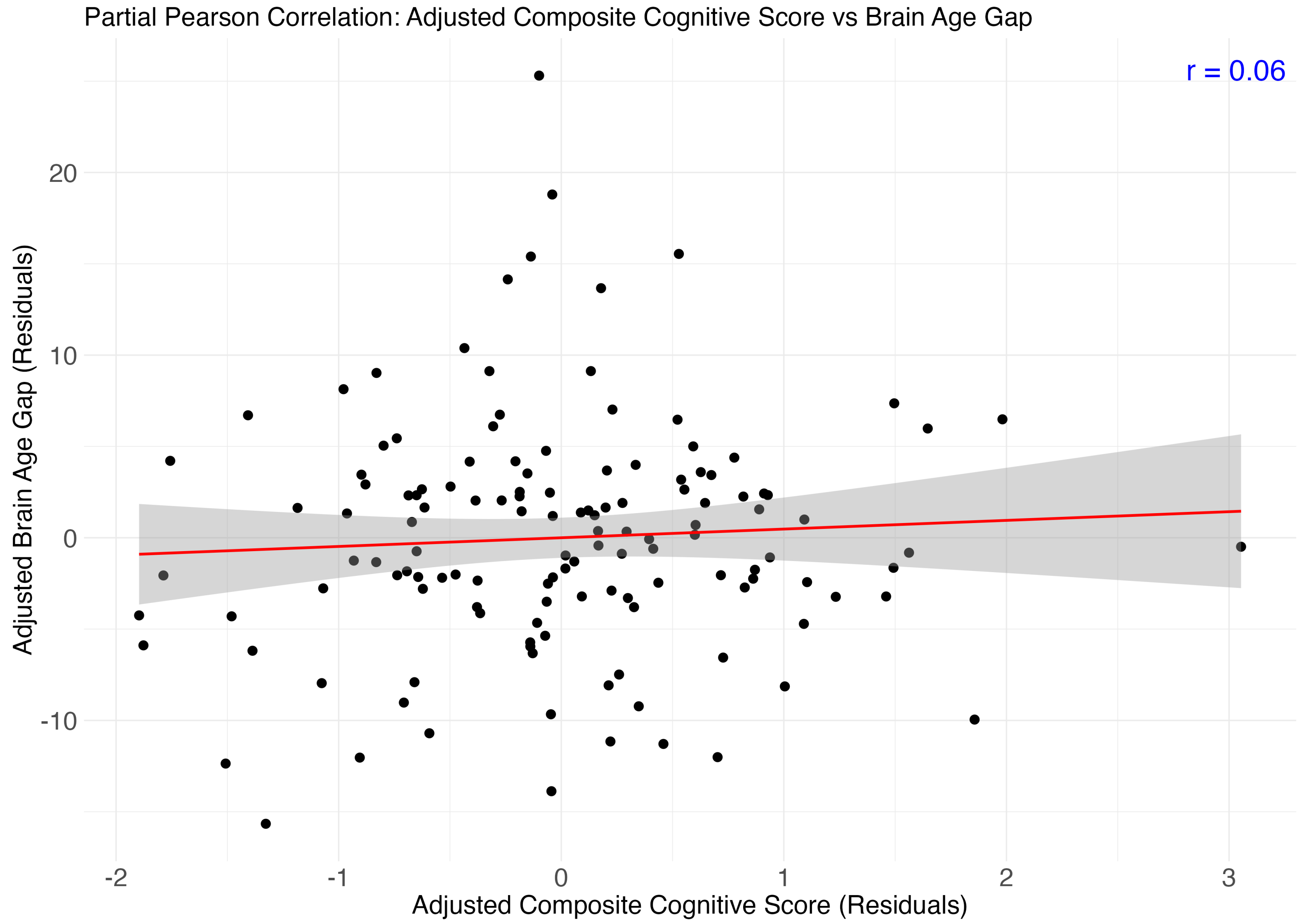
Partial Pearson Correlation: Adjusted Composite Cognitive Score vs Brain Age Gap
r = 0.06
Adjusted Brain Age Gap (Residuals)
20
10
0
-10
-2
-1
0
1
2
3
Adjusted Composite Cognitive Score (Residuals)

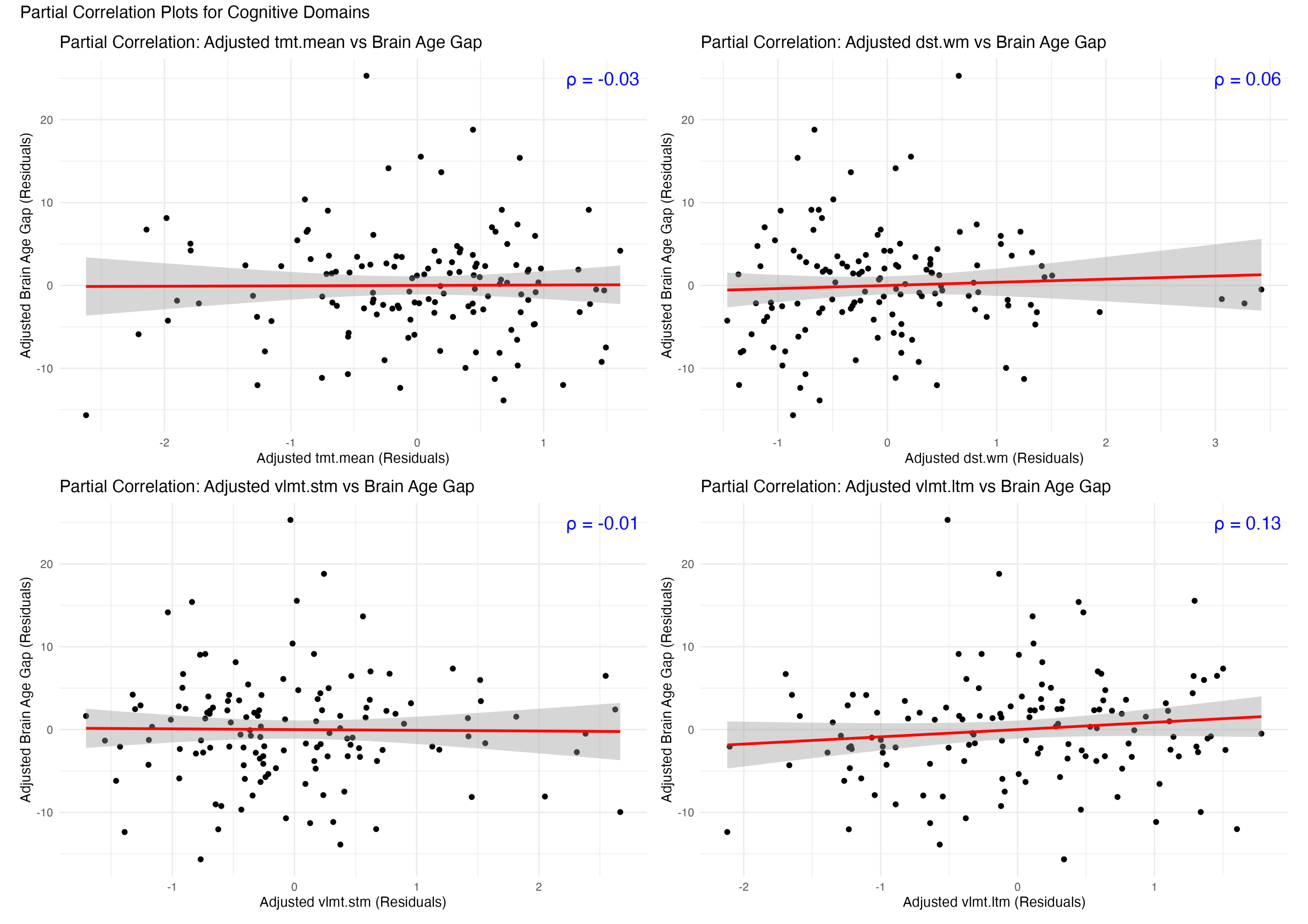
Partial Correlation Plots for Cognitive Domains
Partial Correlation: Adjusted tmt.mean vs Brain Age Gap
ρ = -0.03
Adjusted Brain Age Gap (Residuals)
Adjusted tmt.mean (Residuals)
Partial Correlation: Adjusted dst.wm vs Brain Age Gap
ρ = 0.06
Adjusted Brain Age Gap (Residuals)
Adjusted dst.wm (Residuals)
Partial Correlation: Adjusted vlmt.stm vs Brain Age Gap
ρ = -0.01
Adjusted Brain Age Gap (Residuals)
Adjusted vlmt.stm (Residuals)
Partial Correlation: Adjusted vlmt.ltm vs Brain Age Gap
ρ = 0.13
Adjusted Brain Age Gap (Residuals)
Adjusted vlmt.ltm (Residuals)

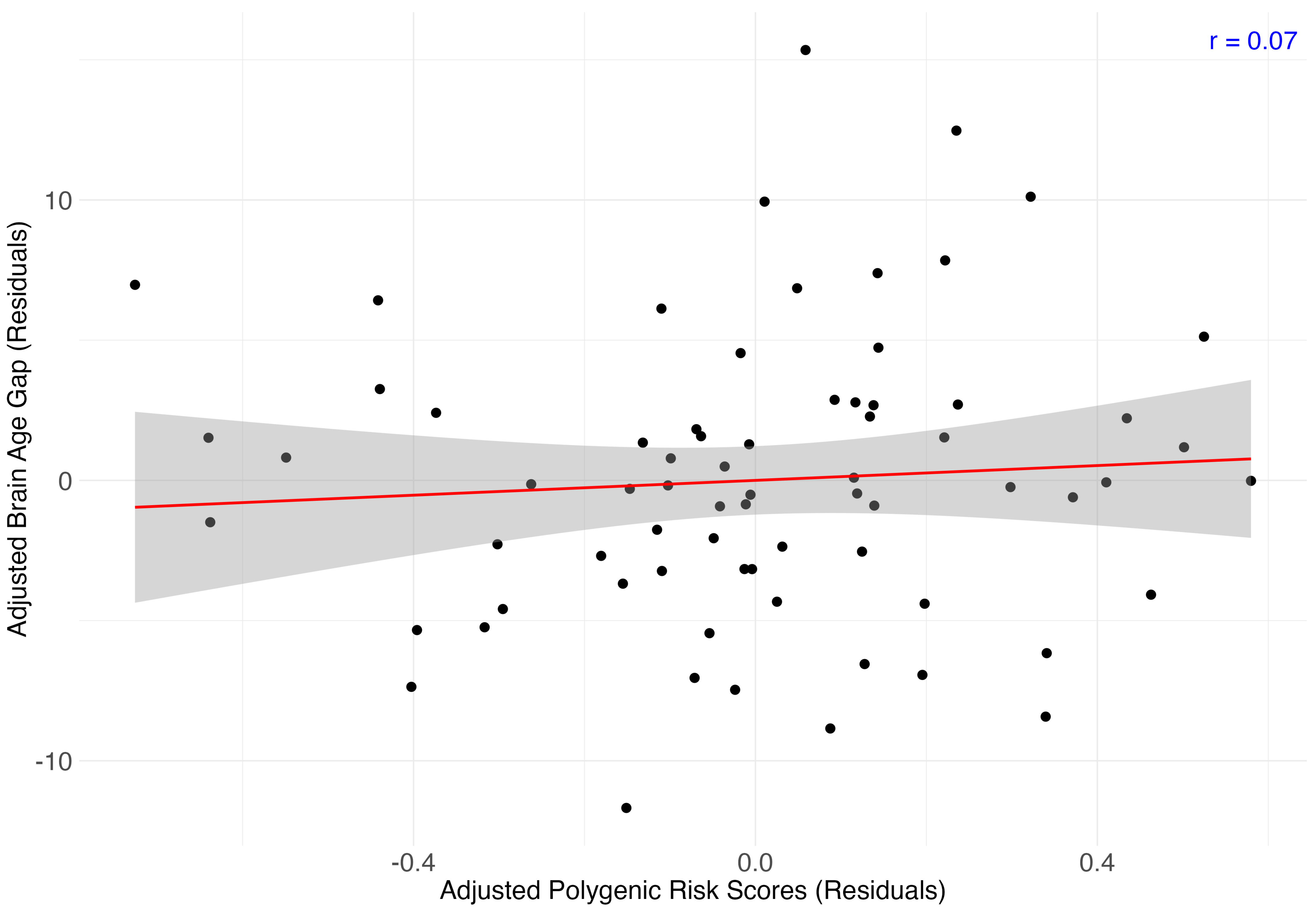
r = 0.07
10
0
-10
-0.4
0.0
0.4
Adjusted Brain Age Gap (Residuals)
Adjusted Polygenic Risk Scores (Residuals)

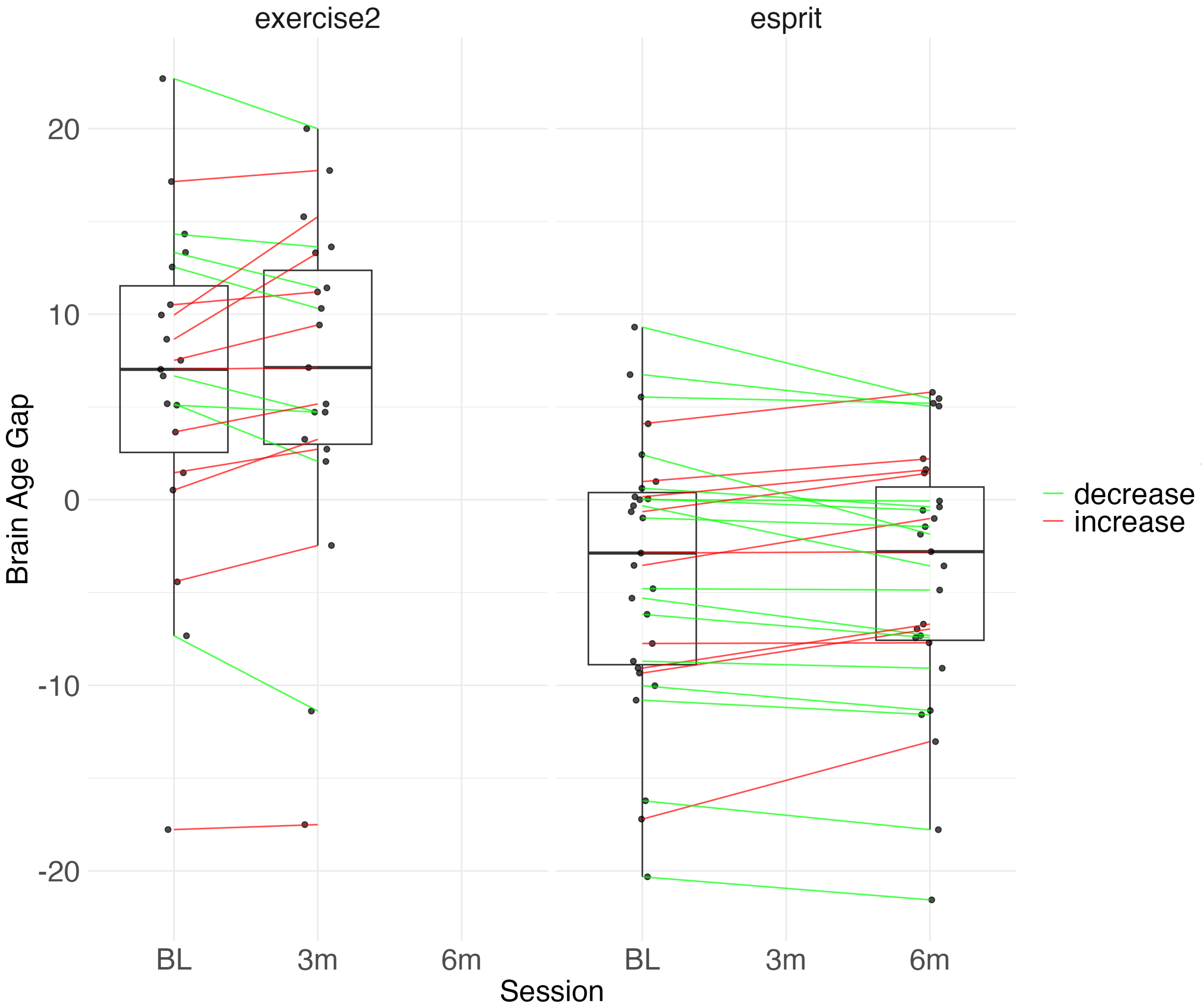
exercise2
esprit
Brain Age Gap
20
10
0
-10
-20
BL
3m
6m
BL
3m
6m
Session
decrease
increase

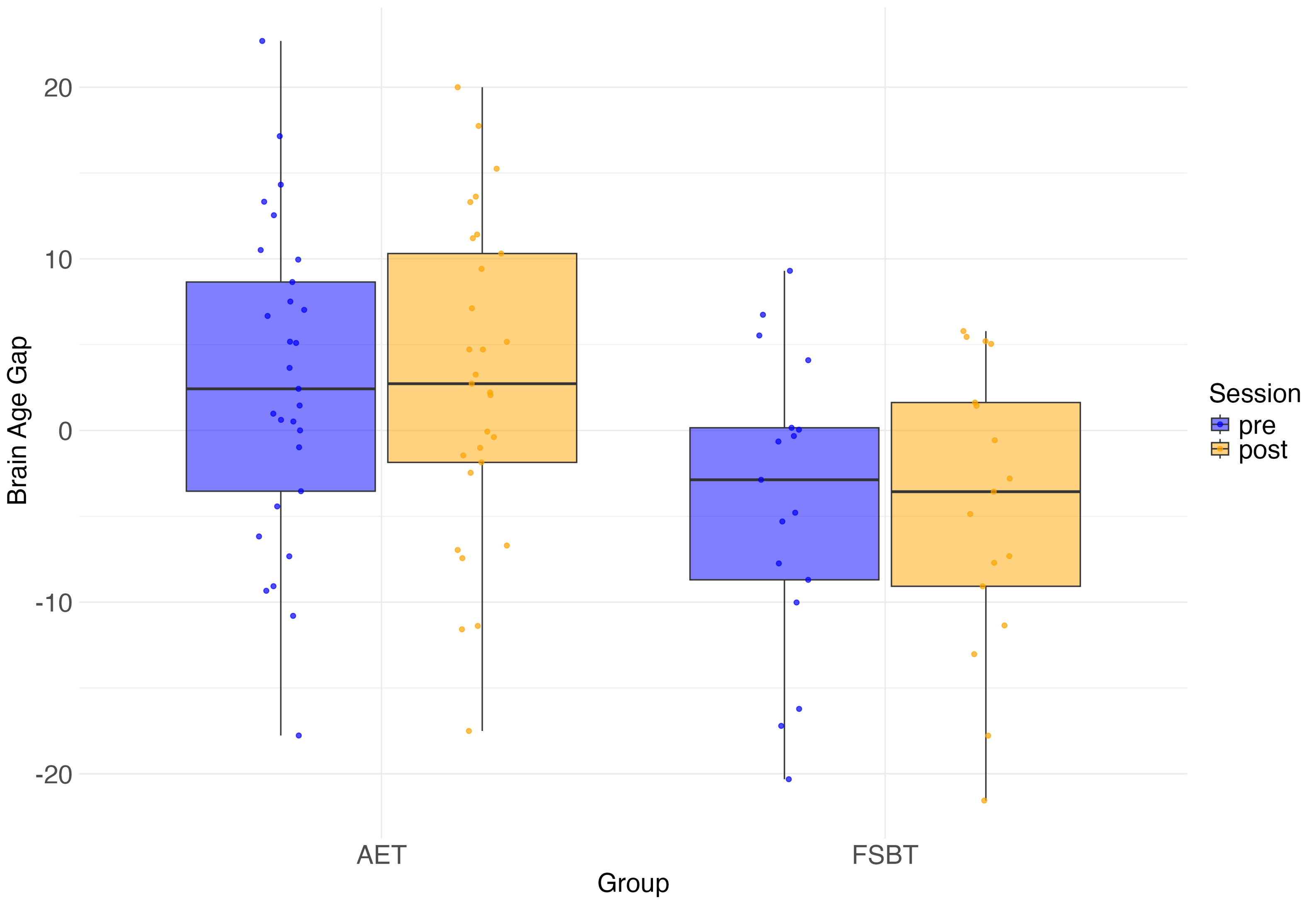
20
10
0
-10
-20
Brain Age Gap
AET
FSBT
Group
Session
pre
post

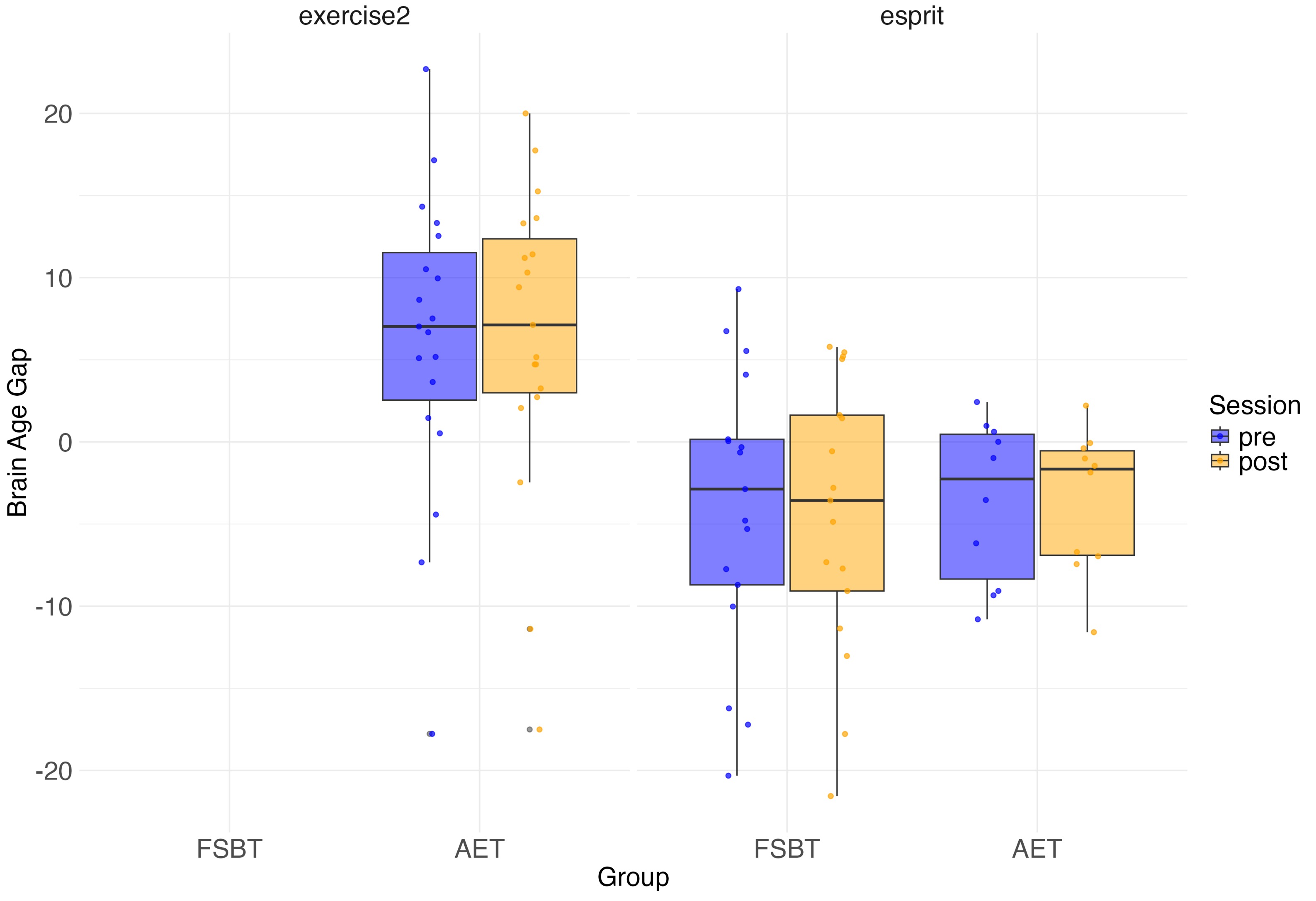
exercise2
esprit
20
10
0
-10
-20
Brain Age Gap
FSBT
AET
FSBT
AET
Group
Session
pre
post

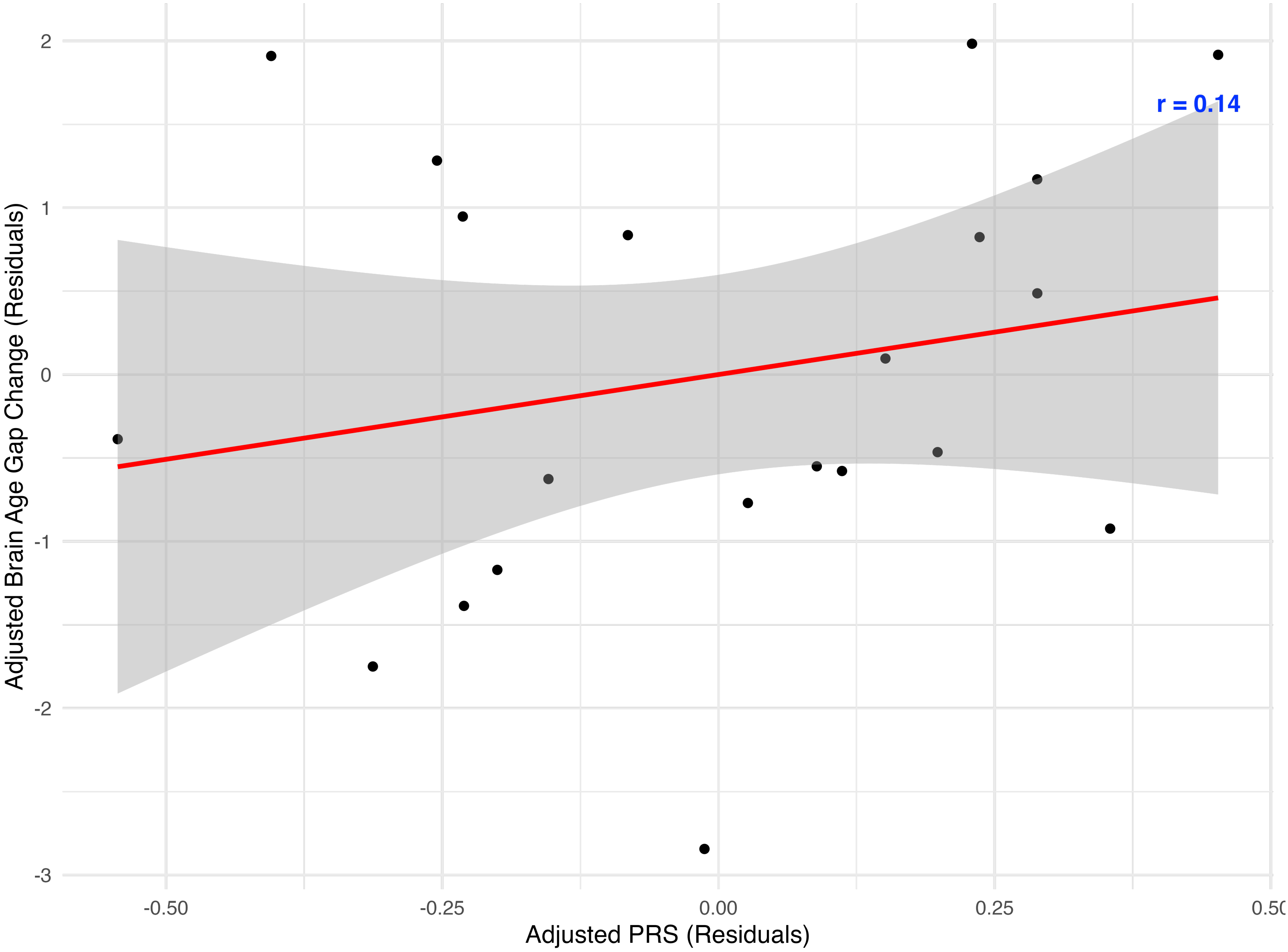

r = 0.14
2
1
0
-1
-2
-3
Adjusted Brain Age Gap Change (Residuals)
-0.50
-0.25
0.00
0.25
0.50
Adjusted PRS (Residuals)
